# Upper airway disease in primary ciliary dyskinesia: Clinical management and factors influencing decision-making, a multicentre analysis

**DOI:** 10.64898/2026.06.08.26354099

**Authors:** Vasiliki Gkatzou, Alexis Campos, Nena Karavasiloglou, Andrea Fernandez-Rodriguez, Mihaela Alexandru, Andreas Anagiotos, Miguel Armengot, Ayse Tana Aslan, I.C.M. Bon, Mieke Boon, Nathalie Isabelle Caversaccio, Suzanne Crowley, Sinan Ahmed D. Dheyauldeen, Eléonore de Garempel de Bressieux, Nagehan Emiralioglu, Ela Erdem Eralp, Yasemin Gokdemir, Eric G. Haarman, Amanda Harris, Isolde Hayn, Hasnaa Ismail-Koch, Bülent Karadag, Oğuzhan Katar, Céline Kempeneers, Dafni Moriki, Ugur Ozcelik, Charlotte O. Pioch, Anne-Lise Poirrier, Johanna Raidt, Ana Reula, Rico N. Rinkel, Tugba Sismanlar Eyuboglu, Stephanie Thee, Panayiotis Yiallouros, Jean-François Papon, EPIC-PCD team, Myrofora Goutaki

## Abstract

**Background:** Upper airway disease is common in primary ciliary dyskinesia (PCD), but management evidence is limited. We aimed to describe management practices and identify factors influencing management decisions.

**Methods:** Using data from the Ear-Nose-Throat (ENT) Prospective International Cohort of patients with PCD (EPIC-PCD) and an ENT-specialist survey across participating centres, we described management practices recorded at routine follow-up. We assessed clinical factors associated with practices via mixed-effects logistic regression models. In a subgroup of patients, we assessed factors associated with initiation or discontinuation of practices.

**Results:** We included 579 patients: median age 15 years, 46% female. Nasal rinsing (54%) and nasal corticosteroids (22%) were most frequently prescribed. Among 466 patients with available data, 47 had grommets (10%) and 42 hearing aids (9%). Nasal corticosteroids and rinsing were more frequently prescribed in patients with polyps (odds ratio [OR] 3.74, 95% confidence interval [CI] 1.80-7.76; OR 3.39, 95% CI 1.37-8.37) or turbinate hypertrophy (OR 1.89, 95% CI 1.03-3.47; OR 2.89, 95% CI 1.55-5.38), and upper airway nebulisation in patients with frequent nasal symptoms (OR 2.86, 95% CI 1.11-7.39). Management practices differed between centres, as seen also by the specialists’ survey responses. In 177 patients with multiple visits, initiation of nasal rinsing was associated with frequent nasal symptoms (OR 3.18, 95% CI 1.24-8.18) and turbinate hypertrophy (OR 3.21, 95% CI 1.20-8.59).

**Conclusion:** Upper airway disease management in PCD varies and is partly guided by symptom burden and clinical findings. This variation across centres highlights the need for care standardisation and PCD-specific management guidelines.

## Introduction

Upper airway disease often is a major and lifelong burden in patients with primary ciliary dyskinesia (PCD), as impaired mucociliary clearance leads to mucus build-up, chronic inflammation, and infections [1–5]. Symptoms usually start in early childhood and continue across the lifetime [4–6]. Acute rhinosinusitis and otitis media with effusion are common [2, 3, 7] and often recur [1, 4, 8]. Over time, these may lead to chronic rhinosinusitis and chronic ear disease, with smell or hearing impairment [2, 3, 8–11]. As a result, patients with PCD require multidisciplinary management addressing also the upper airways as part of standard care [12–15].

Despite the importance of upper airway disease in PCD, evidence to guide its management is limited and remains largely empirical. Existing management recommendations for PCD include only few aspects related to upper airway disease or are based on clinical expertise from single centre studies [12–14, 16–21]. In the absence of management guidelines, clinicians base practices on their clinical experience or adapt them from other chronic airway diseases [13, 22]. It is often unclear when conservative medical practices are sufficient and when surgery is needed. Surgical management remains controversial in PCD, with limited evidence and expert opinions on potential benefits and risks [13, 14, 16–18, 23–28]. However, even for practices that are widely accepted as beneficial, such as nasal saline rinsing [12–15], important aspects remain uncertain such as the recommended frequency, saline volume and concentration used [8, 17, 18, 29]. To be able to support clinical decision-making, it is important to understand how upper airway disease is currently managed in PCD clinical care and which are the most important factors currently influencing management decisions.

In this study, we aimed to comprehensively describe current management practices, identify clinical factors associated with specific disease management practices, and explore the motivation behind clinical decision-making.

## Methods

### Study design and population

We used two data sources: original patient data from the Ear-Nose-Throat (ENT) Prospective International Cohort of patients with PCD (EPIC-PCD) and responses to a survey exploring clinical decision-making among ENT specialists at the participating study centres. This manuscript conforms to the Strengthening the Reporting of Observational studies in Epidemiology (STROBE) statement [30].

#### EPIC-PCD study

EPIC-PCD is an international, multicentre, observational cohort study hosted at the University of Bern (ClinicalTrials.gov identifier: NCT04611516) [31]. It is the first and largest prospective multicentre cohort focusing on upper airway disease in PCD. The study started in February 2020 and follows patients with PCD of all ages during routine follow-up visits. For data collection, the study uses standardised disease-specific tools: the FOLLOW-PCD form and patient questionnaire [32]. We describe the study procedures in detail in the published protocol [31] and Supplementary Methods.

For this study, we used data from 15 participating EPIC-PCD centres (Amsterdam, Ankara (Gazi Hospital), Ankara (Hacettepe Hospital), Athens, Berlin, Bern, Cyprus, Istanbul, Leuven, Liège, Munster, Oslo, Paris, Southampton, and Valencia) across 11 countries. All participating centres obtained ethical approval from local human research ethics committees in accordance with national or local regulations. We obtained informed consent or assent from all patients, or from parents or caregivers of patients aged 14 years or younger, as previously described [31]. Each EPIC-PCD centre established the diagnosis of PCD based on available diagnostic tools according to the European Respiratory Society (ERS) diagnostic guidelines and all participating patients are managed as having PCD [22]. For this manuscript, we classified diagnostic certainty according to the recently published joint ERS and American Thoracic Society (ATS) guidelines (Supplementary Methods) [15].

#### ENT-specialist survey

To gain deeper insights, we developed a survey which aimed to explore decision-making in the management of upper airway disease. We distributed the survey to ENT specialists at all EPIC-PCD participating centres in September 2025. The survey asked about the frequency of recommending commonly prescribed upper airway management practices and related key aspects (Supplementary Methods). Practices included nasal corticosteroids, nasal rinsing, other medical therapies, and surgical practices. We also asked under which circumstances specialists prescribed or recommended these practices, including routine care and exacerbations and which clinical factors they considered most important when making management practice decisions. For children, we asked at which age specialists usually start each management practice. In addition to predefined questions, the survey included free-text fields, where ENT specialists described their motivations and experiences with specific management practices (Supplementary Methods).

### Statistical analysis

We descriptively presented population characteristics and upper airway disease management practices for the total population and stratified by age groups (0–6, 7–12, 13–17, 18–30, 31–50, and ≥51 years), using data entered in the study database that uses the Research Electronic Data Capture (REDCap) software [33] by January 2026. We presented continuous variables using medians and interquartile ranges (IQRs) and categorical variables using counts and proportions.

We examined the association between commonly prescribed upper airway disease management practices and selected clinical factors that influence decisions using multivariable mixed-effects logistic regression models, in patients with available clinical examination and symptoms information at the same visit (Supplementary Figure 1). For each model, we chose factors based on clinical importance and data availability (Supplementary Methods). In all logistic regression models, we included centre as a random effect to account for clustering. Patients with missing information in the variables of interest were excluded from this and the following analyses.

In a subgroup of patients with longitudinal data from repeated follow-up visits, we assessed factors associated with initiation and discontinuation of commonly prescribed practices using mixed-effects logistic regression models (Supplementary Figure 1). At each visit, we recorded whether a patient was prescribed each management practice of interest, namely nasal rinsing, nasal corticosteroids, and upper airway nebulisation (Supplementary Methods). We included the same covariates as in the cross-sectional models and random intercepts for patient and centre to account for repeated observations within individuals and clustering by centre.

We performed all statistical analyses using R software (version 4.4.1; R Foundation for Statistical Computing, Vienna, Austria). We used packages *lme4* for mixed-effects modelling and *ggplot2* for data visualization and used AI-assisted tools to support the development and debugging of code used in the statistical data analysis and visualisation. The results of the logistic regression models were reported as Odds ratios (OR) and 95% confidence intervals (CI).

## Results

### Study population

Out of 597 patients participating in EPIC-PCD, we included 579 patients with available data on upper airway management practices. Included patients had a median age of 15 years (IQR 9–23) and 266 (46%) were female (Table 1). Of the included patients, 395 patients (68%) had confirmed PCD according to ERS/ATS diagnostic guidelines [15] and 350 (60%) had situs solitus.

**Table 1.** Characteristics of EPIC-PCD participants, overall (n=579) and by age group.

|  | **Total**<br>**(n=579)** | **Age 0-6 y**<br>**(n=82)** | **Age 7-12 y**<br>**(n=131)** | **Age 13-17 y**<br>**(n=129)** | **Age 18-30 y**<br>**(n=125)** | **Age 31-50 y**<br>**(n=73)** | **Age >51 y**<br>**(n=39)** |
| --- | --- | --- | --- | --- | --- | --- | --- |
| **Age at assessment, years** | 15 (9-23) | 3 (2-5) | 9 (8-11) | 15 (13-16) | 20 (18-24) | 38 (34-42) | 58 (55-64) |
| **Age at PCD diagnosis, years** | 9 (3-16) | 0.5 (0.1-2) | 5 (2-8) | 10 (7-13) | 15 (11-19) | 34 (29-37) | 46 (43-52) |
| **Female** | 266 (46) | 32 (39) | 57 (44) | 65 (50) | 59 (47) | 31 (42) | 22 (56) |
| **Ethnicity** |  |  |  |  |  |  |  |
| White | 520 (90) | 72 (88) | 120 (92) | 114 (89) | 119 (95) | 60 (82) | 35 (90) |
| Black | 15 (3) | 1 (1) | 3 (2) | 3 (2) | 2 (2) | 5 (7) | 1 (2) |
| Asians | 16 (3) | 3 (3) | 5 (4) | 5 (4) | 3 (2) | 0 (0) | 0 (0) |
| Mixed or unknown | 28 (4) | 6 (8) | 3 (2) | 7 (5) | 1 (1) | 8 (11) | 3 (8) |
| **PCD certainty status<sup>a</sup>** |  |  |  |  |  |  |  |
| Confirmed PCD | 395 (68) | 52 (64) | 85 (65) | 89 (69) | 85 (68) | 57 (78) | 27 (69) |
| Highly likely PCD | 110 (19) | 15 (18) | 20 (15) | 28 (22) | 24 (19) | 13 (18) | 10 (26) |
| Suspected PCD | 74 (13) | 15 (18) | 26 (20) | 12 (9) | 16 (13) | 3 (4) | 2 (5) |
| **Laterality defects** |  |  |  |  |  |  |  |
| Situs inversus | 207 (36) | 37 (45) | 48 (37) | 48 (36) | 51 (40) | 17 (24) | 6 (15) |
| Situs ambiguous | 6 (1) | 0 (0) | 1 (1) | 2 (2) | 1 (1) | 1 (1) | 1 (3) |
| Situs solitus | 350 (60) | 44 (54) | 80 (60) | 78 (61) | 72 (58) | 47 (64) | 29 (74) |
| Unknown | 16 (3) | 1 (1) | 2 (2) | 1 (1) | 1 (1) | 8 (11) | 3 (8) |
| **Cardiovascular malformation present** |  |  |  |  |  |  |  |
| Yes | 53 (9) | 15 (18) | 13 (10) | 11 (9) | 11 (9) | 3 (4) | 0 (0) |
| Unknown | 70 (12) | 6 (7) | 11 (8) | 11 (9) | 16 (13) | 16 (22) | 10 (26) |
| **Bronchiectasis at recruitment** |  |  |  |  |  |  |  |
| Yes | 294 (50) | 9 (11) | 30 (23) | 68 (53) | 96 (77) | 59 (81) | 32 (82) |
| Unknown | 102 (18) | 32 (40) | 30 (23) | 20 (16) | 8 (6) | 7 (10) | 5 (13) |

EPIC-PCD: Ear-Nose-Throat Prospective International Cohort of patients with Primary Ciliary Dyskinesia; HSVM: high-speed video microscopy; IF: immunofluorescence; nNO: nasal nitric oxide; PCD: Primary Ciliary Dyskinesia; TEM: transmission electron microscopy; y: years.
Data is presented as counts (percentages) or median (Interquartile Range). <sup>a</sup>PCD diagnostic certainty status was classified according to the joint European Respiratory Society (ERS)/ American Thoracic Society (ATS) 2025 diagnostic guidelines, as follows: Confirmed PCD: two pathogenic or likely pathogenic variants in a known PCD-associated gene, or a class 1 defect on TEM. Highly likely PCD: class 2 defect on TEM, genetic result concordant with a known genotype–phenotype association, and at least one positive result among nNO, HSVM, or IF. Suspected PCD: not tested with TEM or genetics, or negative results for these tests, with one or more of the following: abnormal beat pattern on HSVM; nasal nitric oxide $\leq 77$ nL/min; pathologic IF staining.

### Upper airway disease management practices

121 (21%) patients reported at least one hospitalisation since their last clinical visit (Table 2). Hospitalisations were most commonly due to lower respiratory infections (n=36, 6%), followed by upper respiratory surgeries (n=27, 5%). Few hospitalisations (≤1%) were due to upper respiratory or other infections. A small proportion of patients (n=30, 5%) reported more than one hospitalisation since their last clinical visit. 218 (38%) patients received at least one course of antibiotics for acute infections, most often for lower respiratory infections (n=154, 27%), while 40 patients (7%) received antibiotics for upper respiratory infections. Penicillin or other β-lactam were the most frequently prescribed antibiotic classes (n=95, 16%). Repeated courses of acute antibiotics were less frequent and when prescribed, they were mainly for lower respiratory infections. Prophylactic antibiotic therapy was reported in 183 patients (32%), most commonly using macrolides (n=134, 23%), and was mainly administered orally. Over half of the patients over the age of 31 years reported use of prophylactic antibiotic therapy (Table 2). 36 (6%) participants reported taking more than one antibiotic prophylactically.

**Table 2.**
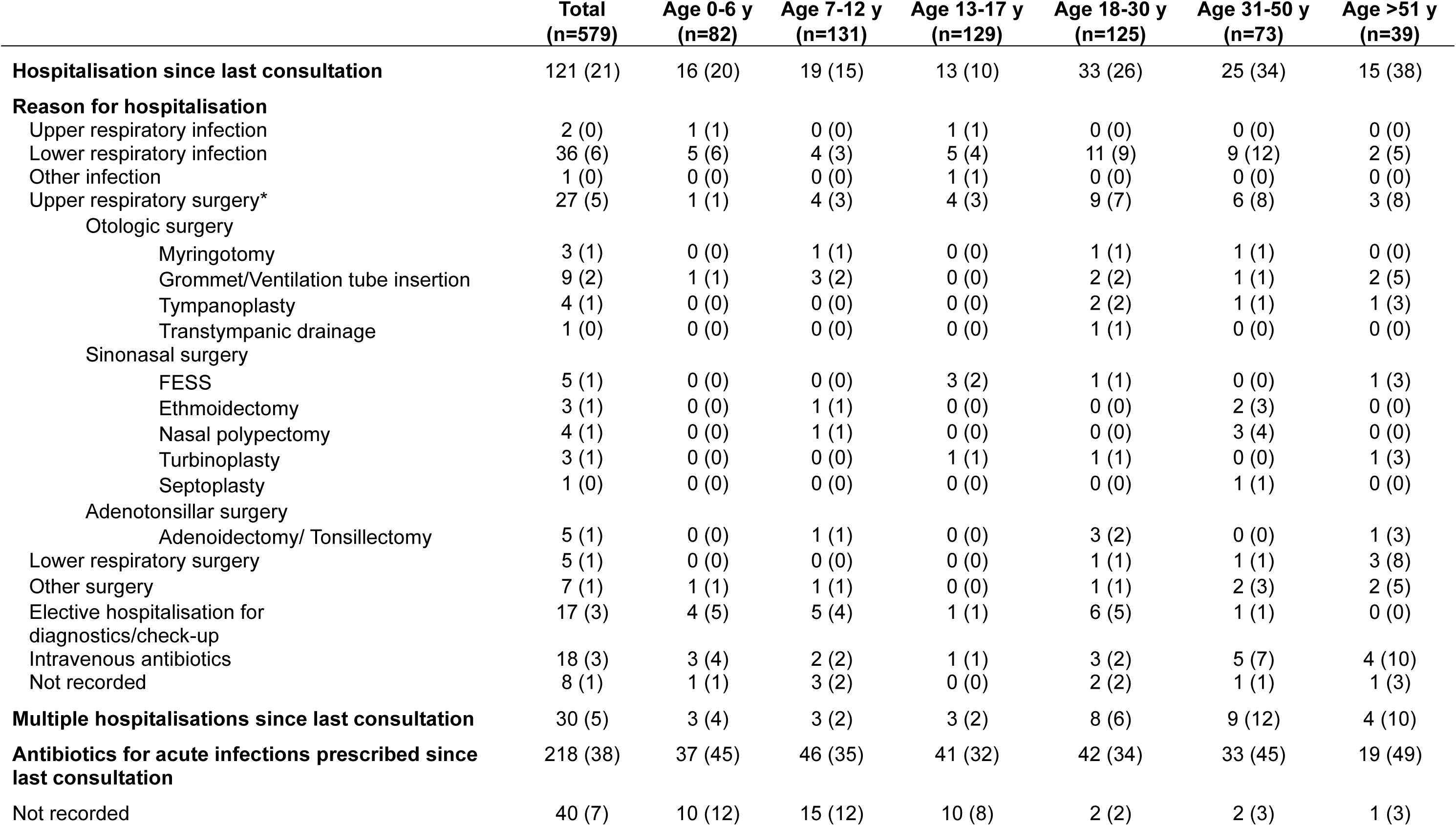
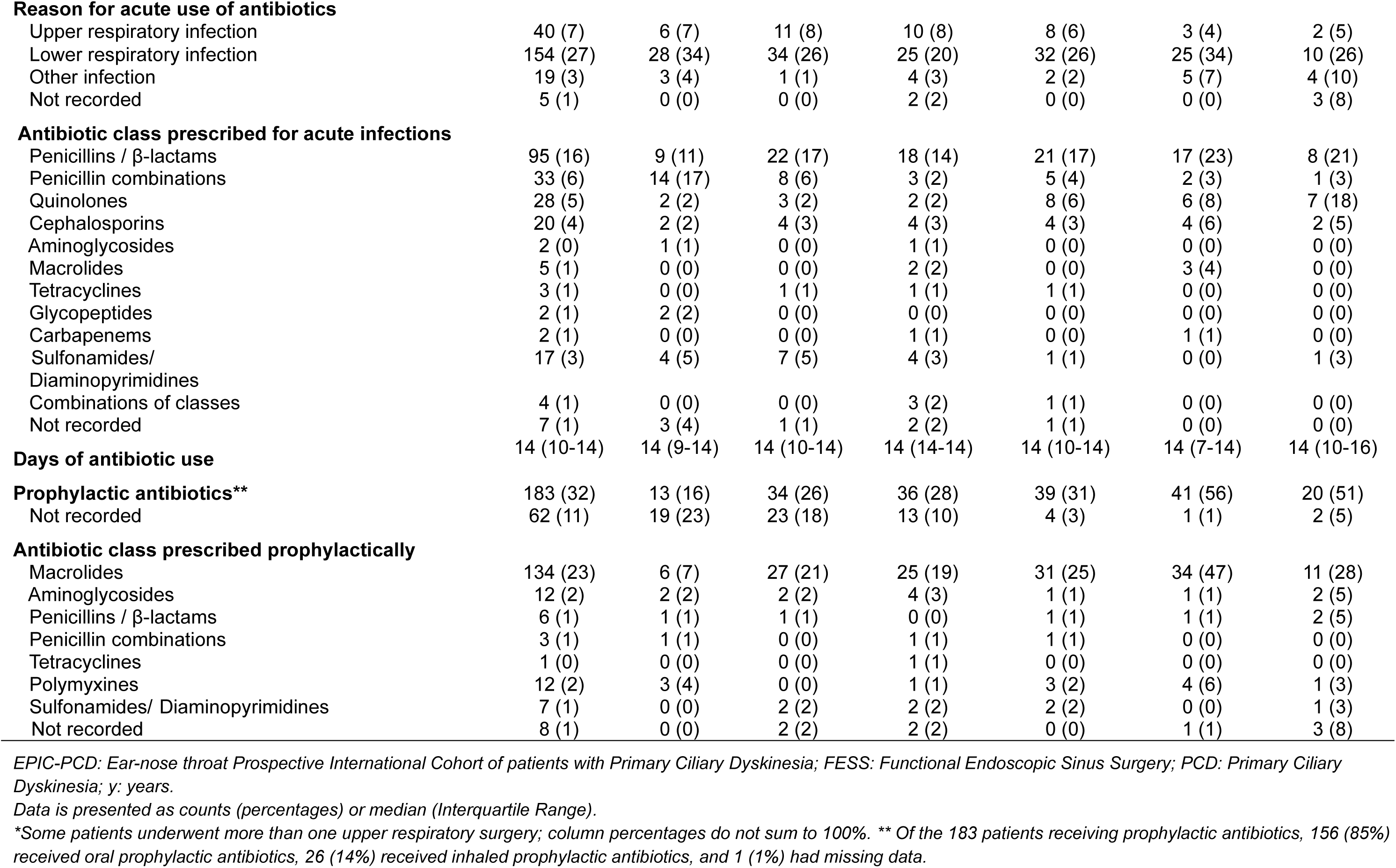
Prevalence of hospitalisations, surgeries and antibiotic use in EPIC-PCD participants, overall (n=579) and by age group.

Regarding routine upper airway management, nasal rinsing was the most frequently prescribed practice (Table 3). 314 patients (54%) were prescribed nasal rinsing, and most were instructed to perform it daily (n=248, 43%). Nasal corticosteroids were prescribed in 125 patients (22%), mostly as continuous year-round therapy. Physicians prescribed less frequently upper airway nebulisation with saline (n=60, 10%) and recommended performing nebulisation almost exclusively daily and most often with hypertonic saline (Table 3). Oral corticosteroids prescription was rare (n=17, 3%) and mainly only during exacerbations (n=11, 2%). Clinical examination data was available for 466 out of 579 patients. Of these, 47 (10%) had grommets and 42 (9%) hearing aids at the time of examination.

**Table 3.**
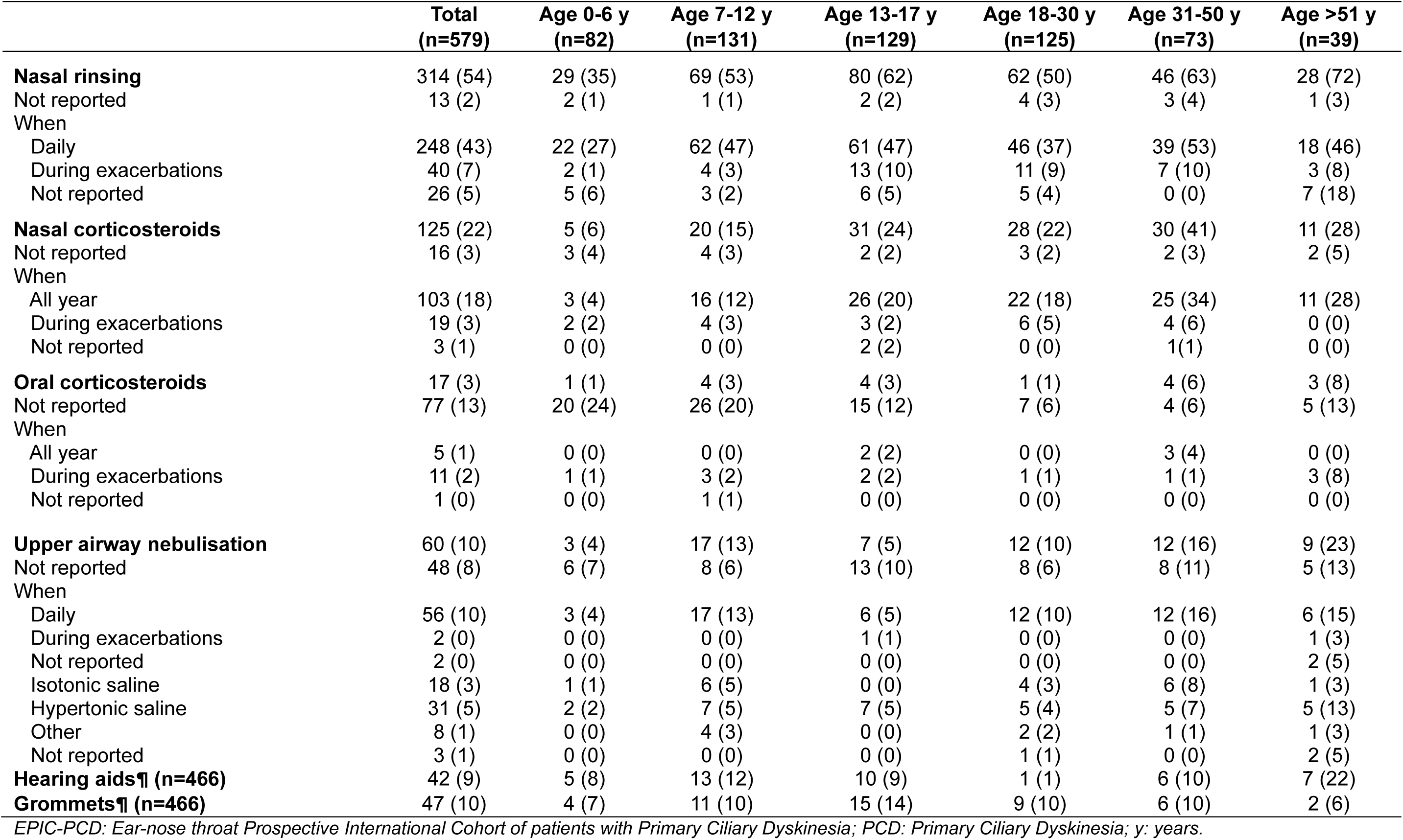
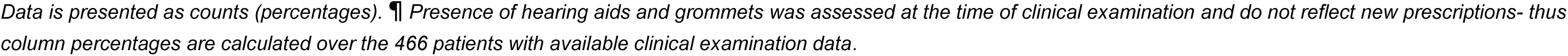
Upper airway disease management practices prescribed in EPIC-PCD participants, overall (n=579) and by age group.

|  | **Total**<br>**(n=579)** | **Age 0-6 y**<br>**(n=82)** | **Age 7-12 y**<br>**(n=131)** | **Age 13-17 y**<br>**(n=129)** | **Age 18-30 y**<br>**(n=125)** | **Age 31-50 y**<br>**(n=73)** | **Age >51 y**<br>**(n=39)** |
| --- | --- | --- | --- | --- | --- | --- | --- |
| **Nasal rinsing** | 314 (54) | 29 (35) | 69 (53) | 80 (62) | 62 (50) | 46 (63) | 28 (72) |
| Not reported | 13 (2) | 2 (1) | 1 (1) | 2 (2) | 4 (3) | 3 (4) | 1 (3) |
| When |  |  |  |  |  |  |  |
| Daily | 248 (43) | 22 (27) | 62 (47) | 61 (47) | 46 (37) | 39 (53) | 18 (46) |
| During exacerbations | 40 (7) | 2 (1) | 4 (3) | 13 (10) | 11 (9) | 7 (10) | 3 (8) |
| Not reported | 26 (5) | 5 (6) | 3 (2) | 6 (5) | 5 (4) | 0 (0) | 7 (18) |
| **Nasal corticosteroids** | 125 (22) | 5 (6) | 20 (15) | 31 (24) | 28 (22) | 30 (41) | 11 (28) |
| Not reported | 16 (3) | 3 (4) | 4 (3) | 2 (2) | 3 (2) | 2 (3) | 2 (5) |
| When |  |  |  |  |  |  |  |
| All year | 103 (18) | 3 (4) | 16 (12) | 26 (20) | 22 (18) | 25 (34) | 11 (28) |
| During exacerbations | 19 (3) | 2 (2) | 4 (3) | 3 (2) | 6 (5) | 4 (6) | 0 (0) |
| Not reported | 3 (1) | 0 (0) | 0 (0) | 2 (2) | 0 (0) | 1 (1) | 0 (0) |
| **Oral corticosteroids** | 17 (3) | 1 (1) | 4 (3) | 4 (3) | 1 (1) | 4 (6) | 3 (8) |
| Not reported | 77 (13) | 20 (24) | 26 (20) | 15 (12) | 7 (6) | 4 (6) | 5 (13) |
| When |  |  |  |  |  |  |  |
| All year | 5 (1) | 0 (0) | 0 (0) | 2 (2) | 0 (0) | 3 (4) | 0 (0) |
| During exacerbations | 11 (2) | 1 (1) | 3 (2) | 2 (2) | 1 (1) | 1 (1) | 3 (8) |
| Not reported | 1 (0) | 0 (0) | 1 (1) | 0 (0) | 0 (0) | 0 (0) | 0 (0) |
| **Upper airway nebulisation** | 60 (10) | 3 (4) | 17 (13) | 7 (5) | 12 (10) | 12 (16) | 9 (23) |
| Not reported | 48 (8) | 6 (7) | 8 (6) | 13 (10) | 8 (6) | 8 (11) | 5 (13) |
| When |  |  |  |  |  |  |  |
| Daily | 56 (10) | 3 (4) | 17 (13) | 6 (5) | 12 (10) | 12 (16) | 6 (15) |
| During exacerbations | 2 (0) | 0 (0) | 0 (0) | 1 (1) | 0 (0) | 0 (0) | 1 (3) |
| Not reported | 2 (0) | 0 (0) | 0 (0) | 0 (0) | 0 (0) | 0 (0) | 2 (5) |
| Isotonic saline | 18 (3) | 1 (1) | 6 (5) | 0 (0) | 4 (3) | 6 (8) | 1 (3) |
| Hypertonic saline | 31 (5) | 2 (2) | 7 (5) | 7 (5) | 5 (4) | 5 (7) | 5 (13) |
| Other | 8 (1) | 0 (0) | 4 (3) | 0 (0) | 2 (2) | 1 (1) | 1 (3) |
| Not reported | 3 (1) | 0 (0) | 0 (0) | 0 (0) | 1 (1) | 0 (0) | 2 (5) |
| **Hearing aids¶ (n=466)** | 42 (9) | 5 (8) | 13 (12) | 10 (9) | 1 (1) | 6 (10) | 7 (22) |
| **Grommets¶ (n=466)** | 47 (10) | 4 (7) | 11 (10) | 15 (14) | 9 (10) | 6 (10) | 2 (6) |

EPIC-PCD: Ear-nose throat Prospective International Cohort of patients with Primary Ciliary Dyskinesia; PCD: Primary Ciliary Dyskinesia; y: years.
*Data is presented as counts (percentages). ¶ Presence of hearing aids and grommets was assessed at the time of clinical examination and do not reflect new prescriptions- thus column percentages are calculated over the 466 patients with available clinical examination data.*

### Factors associated with upper airway management practices

In the regression analyses, we included 466 patients, 222 female (48%) and with median age 15 years (IQR 9–24), who had additional data from ENT examinations (Supplementary tables S1, S2), and on patient-reported symptoms (Supplementary table S3). Prescription of upper airway management practices was associated mainly with specific clinical signs or symptoms rather than sociodemographic factors; however, we found that nasal corticosteroid prescription was more common in men and upper airway nebulisation prescription more common with increased age (Figure 1). Prescription of nasal rinsing was more common in patients with nasal polyps (OR 3.39, 95% CI 1.37–8.37) and turbinate hypertrophy (OR 2.89, 95% CI 1.55–5.38), but we found no signs of association with age, sex, or reported nasal symptom frequency. Prescription of nasal corticosteroids was more common in patients with nasal polyps (OR 3.74, 95% CI 1.80–7.76), inferior turbinate hypertrophy (OR 1.89, 95% CI 1.03–3.47), and frequently reported nasal symptoms (OR 1.74, 95% CI 0.99–3.04). Prescription of upper airway nebulisation was more common in patients reporting frequent nasal symptoms (OR 2.86, 95% CI 1.11–7.39). For grommets, we observed no signs of association with age or sex. We observed substantial centre-level clustering for most upper airway management practices. The intraclass correlation coefficient (ICC) indicated that 45% of the variability in nasal rinsing prescription and 36% of the variability in intranasal corticosteroid prescription were attributable to differences between centres. For upper airway nebulisation, the ICC was particularly high (0.68), suggesting that management practice prescription was largely centre-dependent. In contrast, we saw limited centre-level variability for grommet insertion (ICC 0.08) [34].

**Figure 1:**
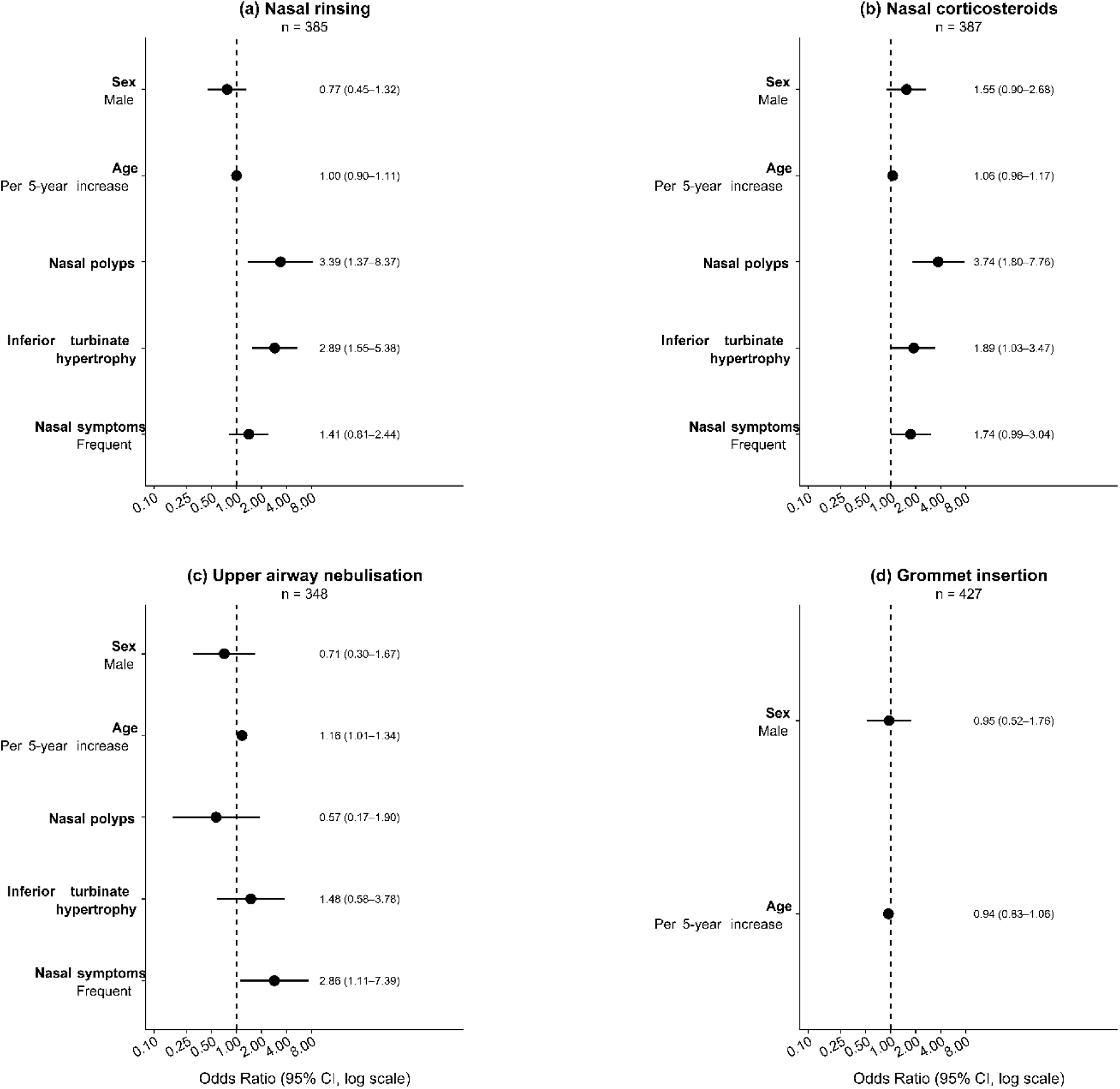
Factors associated with the prescription of (a) nasal rinsing, (b) nasal corticosteroids, (c) upper airway nebulisation and (d) grommet insertion in EPIC-PCD participants. EPIC-PCD: Ear-nose-throat Prospective International Cohort of Patients with Primary Ciliary Dyskinesia. Odds ratios are indicated by circles and 95% confidence intervals by horizontal lines. Reference categories: Sex: Female; Nasal polyps: Absent; Inferior turbinate: Normal; Nasal symptoms: Infrequent.

### Factors associated with initiation or discontinuation of upper airway management practices

In these analyses, we included 177 patients (median age of 17 years (IQR 11–33), 85 female (48%)) with available longitudinal data and a median follow-up of 1.93 years. We explored upper airway disease management practices’ initiation and discontinuation across all available clinical visits. For most patients, management practices were maintained across visits, with 27% changing nasal rinsing practices, 31% nasal corticosteroids practices, and 19% upper airway nebulisation practices (Figure 2). Presence of grommets changed in 8% of patients over time.

**Figure 2.**
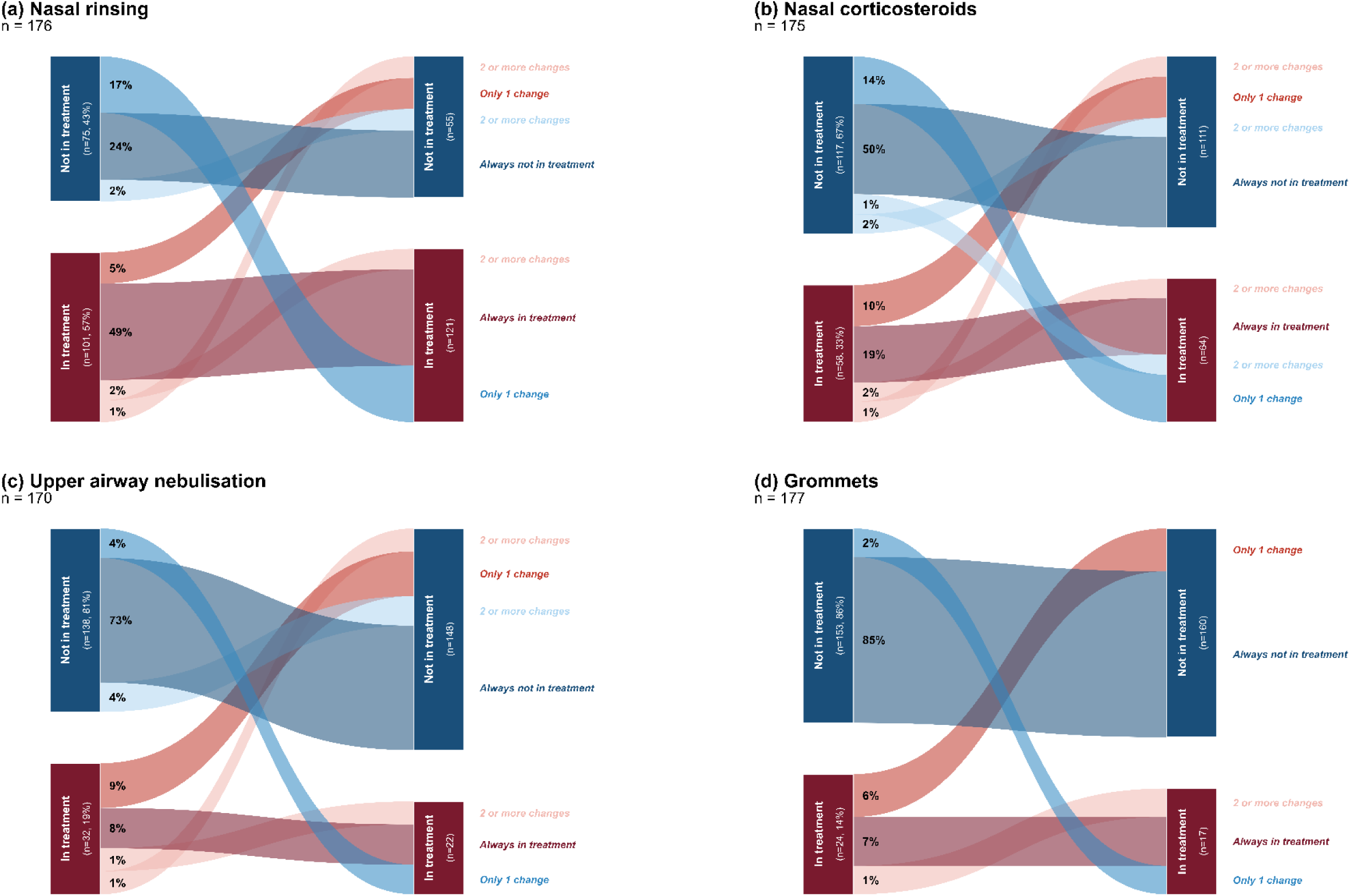
Changes in (a) nasal rinsing, (b) nasal corticosteroids, (c) upper airway nebulisation (d) grommet insertion practices across the available clinical visits over the study period among EPIC-PCD participants. Patients were classified as always following a management practice, always not following a management practice, one change, or ≥2 changes across follow-up. Only patients with ≥1 clinical visits with available data over the study period were included. The median number of visits per patient: 2 (Interquartile range (IQR): 1–2). Percentages are calculated based on the number of patients at baseline.

Initiation of nasal rinsing was associated with frequent patient-reported nasal symptoms (OR 3.18, 95% CI 1.24–8.18) and hypertrophic turbinates at examination (OR 3.21, 95% CI 1.20–8.59) (Figure 3). Initiation of nasal corticosteroids was associated with the presence of nasal polyps (OR 2.40, 95% CI 0.95-6.04). We found no signs of association between initiation of upper airway nebulisation and any studied patient-level clinical factors. The random intercept variance for centre was 0.215 (ICC 0.061), indicating low between-centre variability in nasal rinsing with centre-level differences accounting for 6.1% of the variance in nasal rinsing. We observed no centre effect for nasal corticosteroids initiation, while centre-level variability for nebulisation was moderate (ICC 0.29). Regarding discontinuation of management practices, we observed no signs of association between studied patient-level clinical factors, namely sex, age, and frequency of patient-reported nasal symptoms and any of the studied management practices (Figure 3). Additionally, we did not find between-centre variability in any of the discontinuation models (random intercept variance 0; ICC 0), indicating absence of centre-level clustering in management practice discontinuation decisions.

**Figure 3.**
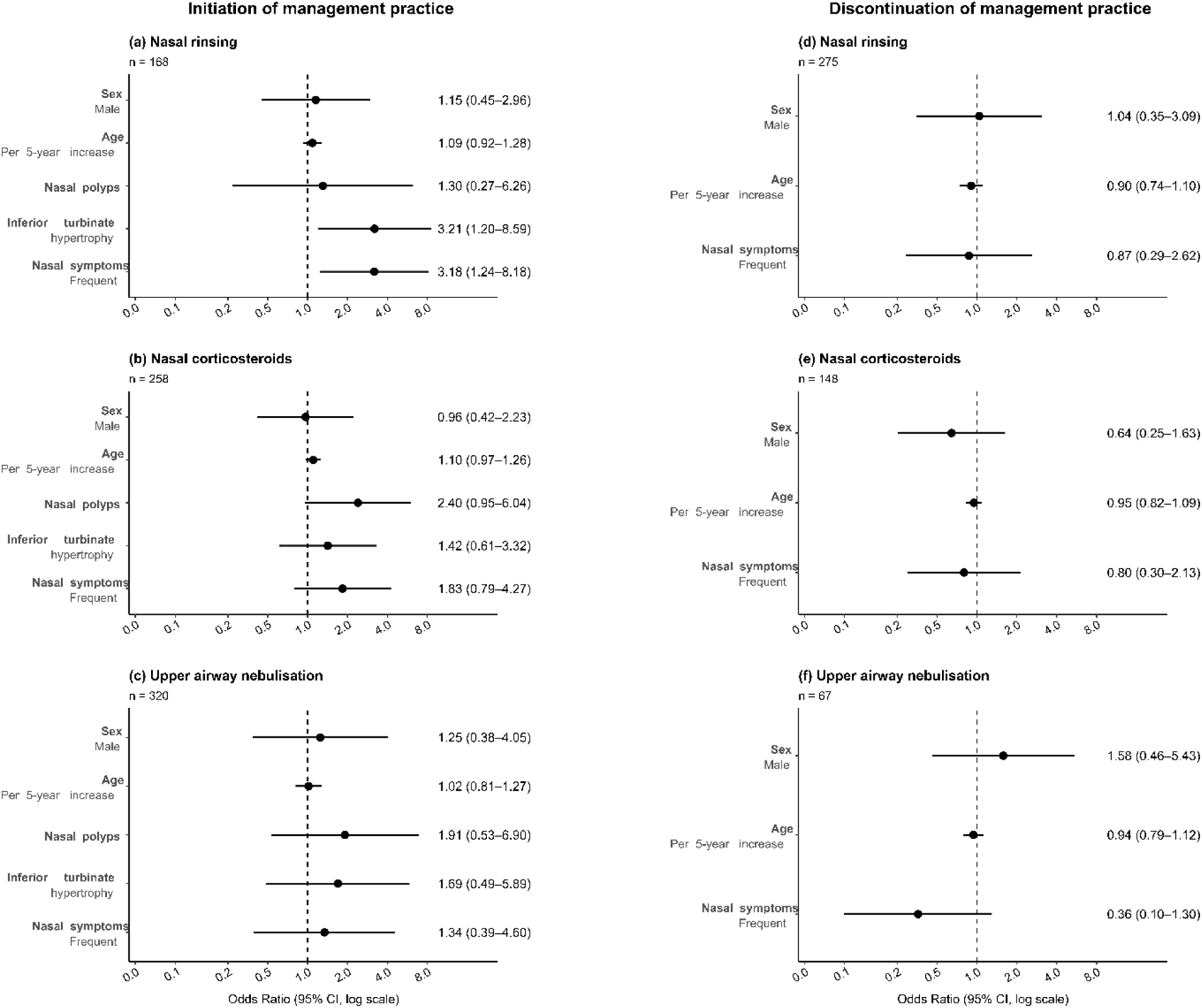
Factors associated with initiation (a,b,c) and discontinuation (d, e, f) of (a,d) nasal rinsing, (b,e) nasal corticosteroids, and (c,f) upper airway nebulisation in EPIC-PCD participants over the study period. EPIC-PCD: Ear-nose-throat Prospective International Cohort of Patients with Primary Ciliary Dyskinesia. Odds ratios are indicated by circles and 95% confidence intervals by horizontal lines. Reference categories: Sex: Female; Nasal polyps: Absent; Inferior turbinate: Normal; Nasal symptoms: Infrequent.

### ENT specialists’ survey results

One ENT specialist from each EPIC-PCD participating centre completed the survey, in total 14 specialists, including three treating only children and 11 treating both children and adults (Supplementary Results). The 15th participating study centre joined the study after circulation of this survey thus no specialist participated. Specialists managing all age groups completed the questions separately for paediatric and adult care. ENT specialists reported nasal rinsing as the most frequently prescribed practice across centres, both during routine care and exacerbations (children: 10/14, 71%; adults: 8/11, 73%) (Figure 4). They prescribed nasal corticosteroids less frequently, with examination findings and patients’ reported symptoms being the main drivers for suggesting these practices (Table 4). Furthermore, they only selectively recommended surgeries (Figure 4, Supplementary Figure 2). In children, age was an important influencing factor, with surgical practices mostly avoided in those below 12 years old (Table 4).

**Figure 4.**
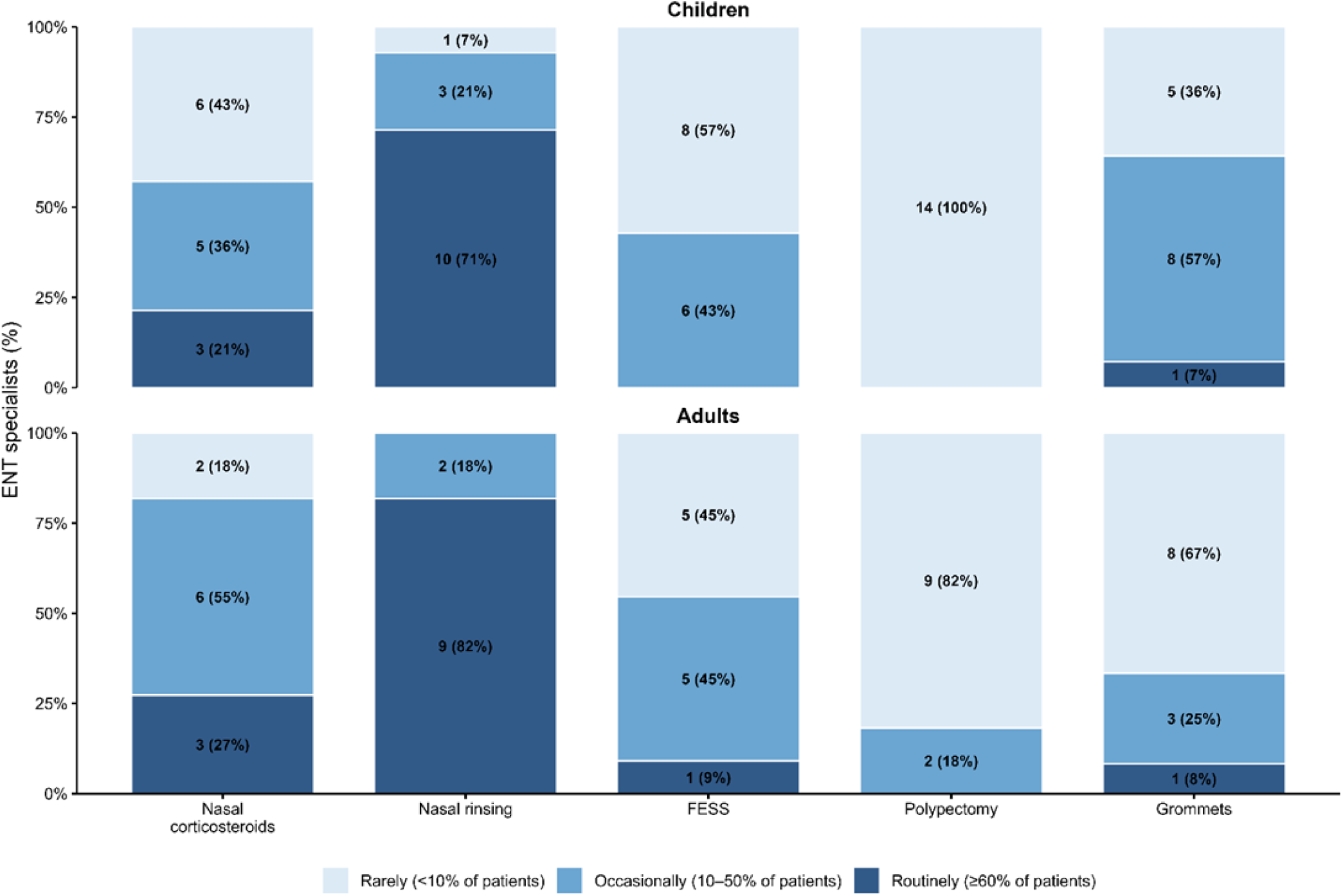
Reported preferred prescription practices of nasal corticosteroids, nasal rinsing, and surgical practices from ENT-specialists at EPIC-PCD participating centres. ENT: Ear-nose-throat; EPIC-PCD: Ear-nose-throat Prospective International Cohort of Patients with Primary Ciliary Dyskinesia; FESS: Functional Endoscopic Sinus Surgery. Bars represent the proportion of ENT specialists prescribing each management practice rarely (<10% of patients), occasionally (10–50% of patients), or routinely (≥60% of patients). Percentages are calculated within each age group, as 3 specialists were treating only children and 11 were treating both children and adults (Children: n = 14; Adults: n = 11). ENT specialists treating both children and adults completed the survey separately for paediatric and adult care. Values inside bars represent the count of respondents and the corresponding percentage.

**Table 4.** Factors influencing recommendation of different upper airway disease management practices reported by ENT specialist at the EPIC-PCD participating centres.

| Category | Children (n=14) |  | Adults (n=11) |  |
| --- | --- | --- | --- | --- |
|  | Decision factors<br>(reported by ≥ 3 ENT-specialists) | Additional considerations | Decision factors<br>(reported by ≥ 3 ENT-specialists) | Additional considerations |
| **Nasal corticosteroids** |  |  |  |  |
| Symptom burden | Nasal obstruction; Nasal discharge | Smell disorder; QoL impairment | Nasal discharge | Nasal obstruction; Smell disorder; QoL impairment |
| Examination findings | Nasal polyps | Turbinate hypertrophy | Nasal polyps | - |
| Comorbidities | - | Allergy; Sleep-related symptoms | - | - |
| Disease severity | - | Complications | - | Complications |
| **Nasal rinsing** |  |  |  |  |
| Symptom burden | Secretions | Congestion/crusting; General symptoms; QoL impairment; Hearing status | - | Secretions; General symptoms; QoL impairment |
| Practical/adherence factors | Ability/convenience/compliance | Education/training; Poor compliance <5 yrs | - | Education/training |
| Age-related factors | - | Age | - | - |
| System-level factors | Socioeconomic status | - | - | - |
|  | Off label/reimbursement issues | - | - | Off label/reimbursement issues |
| **Functional Endoscopic Sinus Surgery (FESS)** |  |  |  |  |
| Examination and imaging findings | Extensive nasal polyposis disrupting sinus drainage | - | Extensive nasal polyposis | - |
| Symptom burden | Facial pressure/pain/headaches | - | - | Facial pressure/pain/headaches |
| Disease severity | Severe clinical features/extreme phenotype | Acute rhinosinusitis complications; Lower airway infections | Severe clinical features/extreme phenotype | Acute rhinosinusitis complications; Lower airway infections |
| Failure of medical therapy | Recurrent exacerbations- no response to treatment | Chronic blockage- not respond to treatment | Recurrent exacerbations- no response to treatment | Chronic blockage- not respond to treatment |
| Age considerations | Not recommended <12 years | - | - | - |
| **Polypectomy** |  |  |  |  |
| Examination findings | - | Massive/high-grade polyps; Total obstruction | - | Massive/high-grade polyps; Total obstruction |
| Symptom burden | - | Patient complaints | - | Nasal obstruction, Severe patient's complaints |
| Disease severity | Extreme phenotype/ Massive-High grade of nasal polyps | Total obstruction; Neoplasm | - | Sinonasal exacerbations; Extreme phenotype; Neoplasm; High grade nasal polyps; Isolated polyps recognised |
|  | Decision factors<br>(reported by ≥ 3 ENT-specialists) | Additional<br>considerations | Decision factors<br>(reported by ≥ 3 ENT-specialists) | Additional<br>considerations |
| Failure of medical therapy | - | Chronic rhinosinusitis unresponsive to treatment | - | Refractory to conservative treatment disease; History of FESS |
| Age considerations | - | Not performed <12 years | - | - |
| General considerations | Generally avoided without FESS | General anaesthesia concerns | - | Patient wish; Generally avoided without FESS; General anaesthesia concerns |
| **Grommets insertion** |  |  |  |  |
| Disease severity | Severe chronic otitis media;<br>Recurrent infections; Hearing loss | Ear surgical history | Severe chronic otitis media;<br>Recurrent infections; Hearing loss | Ear surgical history; pressure |
| Device/adherence factors | - | Inability to tolerate/Low adherence to hearing aids;<br>Parents' request | - | Inability to tolerate/Low adherence to hearing aids;<br>Patient's request |
| Failure of medical therapy | Refractory to conservative treatment otitis media | - | Refractory to conservative treatment otitis media | - |
| Age considerations | Learning problems; Speech acquisition delay; Developmental stage; Age | - | - | - |
| Systemic context | - | Lung status; Limited QoL | - | Lung status; Concern of general anaesthesia; Limited QoL |
| General considerations | - | Trial use; Concern of chronic otorrhea after grommets insertion | - | Trial use; Concern of chronic otorrhea after grommets insertion |

ENT: Ear-nose-throat; EPIC-PCD: Ear-Nose-Throat Prospective International Cohort of patients with Primary Ciliary Dyskinesia; FESS: Functional Endoscopic Sinus Surgery; QoL: Quality of life.
Free-text responses were reviewed and grouped into categories. Decision factors refer to factors reported by ≥3 ENT-specialists who are treating the corresponding age group; less frequently reported factors are reported under additional considerations.

## Discussion

In this large international cohort of patients with PCD, we described comprehensively upper airway disease management in routine clinical care and during exacerbations. Besides infection management with antibiotics, nasal rinsing and corticosteroids were the most commonly prescribed practices and prescription patterns were largely guided by clinical examination findings and symptom burden, with presence of polyps, turbinate hypertrophy, and chronic nasal symptoms often playing the most important role. However, we observed substantial variation between study centres, which was supported by the survey responses from ENT specialists, suggesting that management decisions are not solely determined by patient characteristics but also reflect local practice patterns, availability of therapies, and reimbursement policies. For most patients, upper airway management practices were maintained across visits during the study period.

Nasal rinsing was the most common prescribed management practice in our cohort, prescribed to 54% of patients. This prescription frequency is higher than previously reported estimates in single centre studies in children with PCD or across all ages where it ranged from 3% to 40% [4, 6, 7, 19, 21, 35, 36], but lower than the one reported in adults with PCD in France [8]. Nasal corticosteroids were prescribed to 22% of our patients, similarly to studies in Israel [6] and in Canada [7], but lower than those reported in North American children (50%) [35] and in adults in France (73%) [8]. Upper airway saline nebulisation was used daily by 10% of our patients similarly to previous data from the Swiss PCD registry [37]. Differences in management across centres even within the same country, including in saline and corticosteroid prescription, have been previously reported in Italy [38]. These differences highlight that management remains largely empirical and influenced by local strategies and availability.

Despite the variability in management practices, the reduction in centre variability for management initiation decisions may indicate that clinicians apply more similar criteria when starting new treatments than when maintaining long-term treatment patterns. These findings should, however, be interpreted with caution. The limited range of available management practices may naturally constrain variability in management escalation decisions. It is also possible that important factors of management practice initiation, such as frequency of exacerbations, patient preference and compliance, previous treatment response, or local availability and reimbursement policies, play an important role but these factors could not be included in our analyses due to data unavailability. Upper airway nebulisation with saline showed a more persistent centre effect. Although centre clustering decreased compared with the cross-sectional model, it remained moderate in the initiation analysis and no measured patient-level factors were associated with initiation. This may reflect the fact that nebulisation techniques are more commonly prescribed in specific centres compared to others. On the other hand, no centre-level clustering was observed for discontinuation decisions, and we found no signs of an association with any of the factors studied. Decisions about discontinuation of prescribed practices may therefore depend on other unmeasured factors and in the absence of clear evidence-based guidelines these decisions may remain individualised.

### Strengths and limitations

Our study has several strengths. EPIC-PCD is the first prospective international cohort focused on upper airways disease in PCD and combines clinical examination findings with patient-reported symptoms. The cohort includes a large number of paediatric and adult patients from multiple countries, and we obtained all clinical information during routine clinical visits using standardised FOLLOW-PCD modules, ensuring harmonised data collection across centres. EPIC-PCD has a high response rate across all centres and most invited patients agree to participate since the study is nested in routine clinical care. However, patients with more sinonasal symptoms may be more willing to join when invited. For this study, we excluded patients who did not meet the eligibility criteria, due to lack of management data or the lack of data from clinical examinations and reported symptoms collected at the same visit. We have no reason to believe these exclusions introduced systematic bias and consider them mostly due to organizational aspects at the centres e.g. questionnaires might not have been distributed on some dates. In addition to describing comprehensively upper airway management practices and assessing factors influencing decision-making, the longitudinal design allowed us to examine for the first time management practice initiation and discontinuation over time in a large subsample of patients with available information. However, some limitations should also be considered. Our data reflects prescriptions by specialists rather than the level of adherence by patients. Despite our large sample size, some analyses were limited due to small numbers, particularly for less frequently used management practices, which may have limited our power to detect potential associations or study additional factors. We also lacked data on upper airway exacerbations and on treatment response which contribute to management decisions. Differences in the frequency of follow-up visits between centres may also have influenced the results of longitudinal analyses, which could explain why centre variability was attenuated.

## Conclusion

Upper airway disease in PCD is a frequent and lifelong burden for patients; yet, to date, its management is based mostly on limited options and empirical evidence. In the absence of evidence from upper airway management clinical trials to guide physicians, management decisions largely rely on clinical judgement and are influenced by regional practice patterns, availability, or reimbursement policies. Our findings highlight the pressing need for structured, standardised, multidisciplinary management. Together with other published evidence this study will directly contribute to the ongoing effort to develop an international consensus statement for the medical and surgical management of upper airway disease in children and adults with PCD, which has been initiated in the framework of the BEAT-PCD ERS clinical research collaboration [39].

## Supporting information

Supplementary methods and results

## Data availability

The EPIC-PCD dataset includes individual patient data of people with a rare disease. Although data is pseudonymised, data may still include sensitive information which could potentially lead to identifying patients; therefore, patients were not asked to consent having their data deposited or shared publicly. Upon reasonable request, our datasets for the present study are available from the study principal investigator, Myrofora Goutaki.

## Supplementary material

## Acknowledgements

We thank all people with primary ciliary dyskinesia (PCD) and their families participating in EPIC-PCD and PCD support organisations (especially PCD Family Support Group UK, Association ADCP France, Kartagener Syndrom und Primäre Ciliäre Dyskinesie e.V. Deutschland/Deutschschweiz and Asociación Nacional de Pacientes con Discinesia Ciliar Primaria DCP España/PCD Spain) for their close collaboration. We also thank all researchers of the participating centres involved in enrolment, data collection and data entry who work closely with us (listed below as EPIC-PCD team) and Valérie Schwartz (ISPM, University of Bern) for her valuable research assistance. We are grateful for everyone who contributed to translations of the FOLLOW-PCD questionnaire in Danish, Dutch, Flemish, French, Greek, Norwegian, Spanish, Turkish and Ukrainian. During the preparation of this work, the authors used AI-assisted tools as support for simple writing tasks such as grammar and spell checks.

## EPIC-PCD team (in alphabetical order)

Savvas Achilleos (Nicosia General Hospital, Cyprus), Mihaela Alexandru (AP-HP, France), Andreas Anagiotos (Nicosia General Hospital, Cyprus), Miguel Armengot (La Fe University and Polytechnic Hospital, Spain), Orhan Asya (Marmara University, Turkey), Ayse Tana Aslan (Gazi University, Türkiye), Achim Georg Beule (University of Münster, Germany), I.C.M. Bon (Vrije Universiteit, the Netherlands), Romane Bonhiver (University Hospital of Liège, Belgium), Mieke Boon (University Hospital Leuven, Belgium), Noëmie Bricmont (University Hospital of Liège, Belgium), Andrea Burgess (University Hospital of Southampton, UK), Doriane Calmes (University Hospital of Liège, Belgium), Alexis Campos (AP-HP, France), Nathalie Isabelle Caversaccio (University Hospital of Bern), Victoria Collard (University Hospital of Liège, Belgium), Suzanne Crowley (Oslo University Hospital, Norway), Erika Defgnee (University Hospital of Liège, Belgium), Sinan Ahmed D. Dheyauldeen (Oslo University Hospital, Norway), Konstantinos Douros (National and Kapodistrian University of Athens, Greece), Nagehan Emiralioglu (Hacettepe University, Turkey), Ela Erdem Eralp (Marmara University, Turkey), Almala Pınar Ergenekon (Marmara University, Turkey), Andrea Fernandez-Rodriguez (University of Bern, Switzerland), Nathalie Feyaerts (University Hospital Leuven, Belgium), Vasiliki Gkatzou (University of Bern, Switzerland), Yasemin Gokdemir (Marmara University, Turkey), Myrofora Goutaki (University of Bern, Switzerland), Nor el houda Guenaoui (AP-HP, France), Eric G. Haarman (Vrije Universiteit Amsterdam, The Netherlands), Amanda Harris (University of Southampton, UK), Isolde Hayn (Charité-Universitätsmedizin Berlin, Germany), Simone Helms (University of Münster, Germany), Turkey), Isabel Ibáñez (La Fe University and Polytechnic Hospital, Spain), Hasnaa Ismail Koch (University of Southampton, UK), Seyda Karabulut (Marmara University, Turkey), Bülent Karadag (Marmara University, Turkey), Nena Karavasiloglou (University of Bern, Switzerland), Oğuzhan Katar (Hacettepe University, Turkey), Céline Kempeneers (University Hospital of Liège, Belgium), Ammar El Kheir (University Hospital of Liège, Belgium), Panayiotis Kouis (University of Cyprus, Cyprus), Nilgun Kula (Gazi University, Türkiye), Oğuz Kuşçu (Hacettepe University, Turkey), Philipp Latzin (University of Bern, Switzerland), Natalie Lorent (University Hospital Leuven, Belgium), Jane S. Lucas (University of Southampton, UK), Dafni Moriki (National and Kapodistrian University of Athens, Greece), Loretta Müller (University of Bern, Switzerland), Noelia Muñoz (La Fe University and Polytechnic Hospital, Spain), Heymut Omran (University of Münster, Germany), Ugur Ozcelik (Hacettepe University, Turkey), Jean-François Papon (AP-HP, France), Clara Pauly (University Hospital of Liège, Belgium), Sevgi Pekcan (Necmettin Erbakan University, Turkey), Charlotte O. Pioch (Charité-Universitätsmedizin Berlin, Germany), Anne-Lise Poirrier (University Hospital of Liège, Belgium), Johanna Raidt (University of Münster, Germany), Louis Reichenheim (Charité-Universitätsmedizin Berlin, Germany), Ana Reula (CEU-Cardenal Herrera University, Spain), Rico N. Rinkel (Vrije Universiteit Amsterdam, the Netherlands), Suat Savaş (Necmettin Erbakan University, Turkey), Andre Schramm (University of Münster, Germany), Tugba Sismanlar Eyuboglu (Gazi University, Türkiye), Catherine Sondag (University Hospital of Liège, Belgium), Janette Tattersall-Wong (Charité-Universitätsmedizin Berlin, Germany), Stephanie Thee (Charité-Universitätsmedizin Berlin, Germany), Ruth Urbantat (Charité-Universitätsmedizin Berlin, Germany), Panayiotis Yiallouros (University of Cyprus, Cyprus), Cansu Yılmaz Yegit (Marmara University, Turkey)

## Author contributions

M. Goutaki developed the concept and design of the study. M. Goutaki and V. Gkatzou managed the study. V. Gkatzou cleaned, standardised the data and performed statistical analyses, supervised by M. Goutaki and N. Karavasiloglou. V. Gkatzou, N. Karavasiloglou and M. Goutaki drafted the manuscript. All authors commented and revised the manuscript and take full responsibility for the content of this publication.

