## Supplementary methods and results for "Upper airway disease in primary ciliary dyskinesia: Clinical management and factors influencing decision-making, a multicentre analysis"

**EPIC-PCD study methods**

EPIC-PCD study procedures are fully embedded in standard clinical care and do not require any additional study-specific interventions [1]. Ear Nose and Throat (ENT) specialists perform routine ENT consultations at each participating centre. Depending on local protocols and patients' needs, consultations may include nasal endoscopy or anterior rhinoscopy, audiometry, and tympanometry. ENT specialists record findings from these examinations in a standardised way using the ENT examination module of the FOLLOW-PCD form [2]. At each consultation, they also collect information on prescribed management and treatment. Patients report hospitalisations and antibiotic use for acute infections since the last visit, as well as the use of maintenance antibiotics or physiotherapy. Specialists record these data, together with any new prescriptions related to upper airway management, using the "Hospitalisations and Treatment" module of the FOLLOW-PCD form [2]. They record upper airway management practices including the prescription of nasal corticosteroids, nasal rinsing, nebulisation, and oral corticosteroids. We also collect demographic information, including age, sex, ethnicity, and baseline medical history retrospectively from medical electronic records, including laterality defects, cardiovascular malformations, bronchiectasis at study recruitment, and other concomitant diseases.

To collect patient self-reported symptoms, we use the disease-specific FOLLOW-PCD questionnaire (version 1.0) [2]. The questionnaire is available in three age-specific versions: adults, adolescents aged 14–17 years, and parents or caregivers of children with PCD younger than 14 years. Participating centres use versions translated into their local languages. The questionnaire asks about the type and frequency of sinonasal and otologic symptoms during the past three months. Patients report symptom frequency using a five-point Likert scale (daily, often, sometimes, rarely, and never), with "unknown" as an additional option. Patients complete the questionnaire on the same day as, or within 14 days of the ENT clinical examination. The study stores all collected data in a central database using Research Electronic Data Capture (REDCap) software [3].

Diagnostic certainty was classified according to the joint European Respiratory Society (ERS) and American Thoracic Society (ATS) 2025 diagnostic guidelines [4]. Patients were classified as confirmed PCD if they had two pathogenic or likely pathogenic variants in a known PCD-associated gene, or a class 1 defect identified on transmission electron microscopy (TEM).

Patients were classified as highly likely PCD if they had a class 2 defect on TEM, a genetic result concordant with a known genotype–phenotype association, and at least one positive result among nasal nitric oxide (nNO), high-speed video microscopy (HSVM), or immunofluorescence (IF). Patients were classified as suspected PCD if they had not been tested with TEM or genetics, or had negative results for these tests, but had one or more of the following: an abnormal beat pattern on HSVM; nNO ≤77 nL/min; or pathologic IF staining. All patients included in this analysis were treated as having PCD at their collaborating centre, regardless of certainty class.

**ENT-specialists survey**

To gain deeper insights, we developed a survey which aimed to explore decision-making in the management of upper airway disease. The survey was distributed online via Microsoft Forms to one ENT specialist per participating centre in September 2025. In total 14 specialists, including 3 treating only children and 11 treating both children and adults (Supplementary Results) responded. The 15th participating study centre joined the study after circulation of this survey thus no specialist participated. Reminder emails were sent to non-responding centres to maximise response rates. The survey took approximately 20–30 minutes to complete. The survey was organised into separate sections for each management practice, covering paediatric and adult care independently. We used a combination of multiple-choice questions and free-text fields. Multiple-choice questions asked about prescription frequency, clinical context (routine care or exacerbations), preferred treatment schemes including dose, frequency, and duration, and the age at which each management practice is typically initiated in children. Free-text fields invited specialists to elaborate on their clinical reasoning, describe factors that would lead them to modify the way a management practice was prescribed, and share any uncertainties or challenges related to each practice. The full survey is available at the end of this Supplementary Methods document.

**Definition of outcomes and covariate selection for statistical analysis models**

Definition of outcomes and covariate selection

For our analysis, we included available data on upper airway management and data from ENT examinations and completed symptom questionnaires on the same day as, or within 14 days of the ENT examination. We selected nasal corticosteroids, nasal rinsing, nebulisation, and ventilation tube insertion as the management practices of interest, based on their frequent use and clinical relevance. We considered several covariates, based on clinical relevance and data availability. Regarding patients' demographic information, we included age (continuous, in years), sex (female/male) and participating centre. From clinical ENT examinations, we considered the responses recorded by specialists related to the presence of nasal polyps (yes/no) and nasal turbinate status (normal/hypertrophic). Nasal turbinate status was originally recorded across four categories: normal, hypertrophic, atrophic, and not assessed. We excluded patients with atrophic turbinates (n = 7) because of the small number of cases and because this finding was not considered clinically relevant for treatment decisions. For both variables, nasal polyps and nasal turbinate status, we recoded responses recorded as “not assessed” or “unknown” as missing.

We also considered patient-reported symptom data collected via the disease-specific FOLLOW-PCD questionnaire at the visit or within 14 days [2]. The questionnaire inquired about the frequency of chronic nasal symptoms including nasal obstruction, rhinorrhoea, sneezing, and loss of smell experienced during the past three months. Patients recorded their responses to these questions on a five-point scale (never / rarely / sometimes / often / daily). Parents or caregivers of children with PCD younger than 14 years responded on the child's behalf, ensuring that symptom frequency was captured even in younger age groups unable to self-report. For the purpose of this analysis, the five response categories were recoded into a binary variable to improve analytical clarity and statistical power. Responses of never, rarely, and sometimes were grouped into the “infrequent” category, reflecting the absence of a meaningful symptom burden, while responses of often and daily were grouped into the “frequent” category, reflecting the presence of regular symptoms. Responses recorded as “not assessed” or “unknown,” were coded as missing. This recoded binary variable was used in all subsequent statistical models and descriptive results. We used the same classification approach for all patient-reported symptom variables we presented in Supplementary Table 3.

Statistical analysis considerations

We examined the association between commonly prescribed upper airway disease management practices and selected factors using multivariable mixed-effects logistic regression models, in patients with available clinical examination and symptoms information at the same visit or within 14 days (Supplementary Figure 1). For each model, we chose factors based on clinical importance and data availability, which we included in the models as described above. In all logistic regression models, we included centre as a random effect to account for clustering. Patients with missing information in the variables of interest were excluded from this and the following analyses.

For grommets insertion, a different modelling strategy was used because the study records presence of grommets at the clinical ENT examination rather than a new management decision. Thus, clinical findings recorded at the same visit may reflect consequences rather than decision triggers. Therefore, models for grommets included only age and sex as fixed effects and centre as a random effect.

In a subgroup of patients with longitudinal data from repeated follow-up visits, we assessed factors associated with initiation and discontinuation of commonly used practices using mixed-effects logistic regression models. When assessing practice initiation, we included only patients who were not prescribed the practice at their first available visit. The outcome was prescription of each management practice at a subsequent visit (hereafter called initiation). For discontinuation models, we included patients who were prescribed the practices at their first available visit. The outcome was not prescribing the management practice at a subsequent visit (hereafter called discontinuation). After initiation or discontinuation of a practice, follow-up was censored and no further events were considered for this patient. We modelled the first initiation or discontinuation event only. In the initiation models, we included the same covariates as in the cross-sectional models (as described above) and random intercepts for patient and centre to account for repeated observations within individuals and clustering by centre. In the discontinuation models, we included covariates that were clinically relevant to stopping a management practice namely sex, age, and frequency of patient-reported nasal symptoms (classified as described above) and not covariates which are unlikely to change rapidly and are less directly related to short-term management practice decisions.

### ENT specialists' survey

### General Questions

1. At which centre do you work?

*(Please select only one from the options below.)*

☐ Amsterdam

☐ Ankara

☐ Berlin

☐ Bern

☐ Nicosia

☐ Istanbul

☐ Leuven

☐ Liege

☐ Münster

☐ Oslo

☐ Paris

☐ Southampton

☐ Valencia

2. What are the age groups of PCD patients that you treat?

*(Please select only one from the options below.)*

☐ Only children

☐ Only adults

☐ Both children and adults

### Medication prescription practices

### Nasal corticosteroids prescription in adults

3. How often do you prescribe nasal corticosteroids to adults with PCD at your centre?

☐ Rarely (in less than 10% of patients)

☐ In some cases (in about 10-40% of patients)

☐ In about half of the cases (in about 50% of patients)

☐ In the majority of cases (in about 60-90% of patients)

☐ Nearly in all cases (in more than 90% of patients)

4. In which of the following cases, do you prescribe nasal corticosteroids to adults with PCD at your centre?

☐ Only during exacerbations

☐ Only during routine care

☐ During both routine care and exacerbations

*The following question: 5, will appear only if the answers in question:4 are: "Only during routine care", "During both routine care and exacerbations")*

5. Please describe in detail the three most important factors (e.g. specific symptoms or signs seen during examination) that make you prescribe nasal corticosteroids to adults with PCD at your centre, in routine care.

*(Free text answer)*

*The following question: 6, will appear only if the answers in question:4 are: "Only during routine care", "During both routine care and exacerbations")*

6. During routine care what is the usual therapeutic scheme of nasal corticosteroids you prescribe to adults with PCD at your centre?

*(Free text answer– Please include details such as the daily dose, frequency, duration of treatment scheme and any other relevant instructions)*

7. Are there any factors that make you **shorten** the usual duration or daily dose of nasal corticosteroids treatment to adults with PCD at your centre?

*(Free text answer - Please describe any clinical or patient-related factors that influence your decision to shorten the usual treatment duration/ daily dose.)*

8. Are there any factors that make you **increase** the usual duration or daily dose of nasal corticosteroids treatment to adults with PCD at your centre?

*(Free text answer - Please describe any clinical or patient-related factors that influence your decision to increase usual treatment duration/or daily dose.)*

*The following question: 9, will appear only if the answers in question:4 are: "Only during exacerbations", "During both routine care and exacerbations")*

9. During exacerbations, what is the usual therapeutic scheme of nasal corticosteroids you prescribe to adults with PCD at your centre?

*(Free text answer– Please include details such as the daily dose, frequency, duration of treatment scheme and any other relevant instructions)*

10. Do you have any additional comments or thoughts on the prescription of nasal corticosteroids to adults with PCD?

(*Free text answer – Please include any thoughts on clinical experience, challenges, preferences, or uncertainties related to their use.)*

### Nasal Corticosteroids prescription in children

(These questions will appear only if they treat children in their centre)

11. How often do you prescribe nasal corticosteroids to children with PCD at your centre?

☐ Rarely (in less than 10% of patients)

☐ In some cases (in about 10-40% of patients)

☐ In about half of the cases (in about 50% of patients)

☐ In the majority of cases (in about 60-90% of patients)

☐ Nearly in all cases (in more than 90% of patients)

12. From what age do you prescribe nasal corticosteroids to children with PCD at your centre?

☐ 2 – 5 years

☐ 6 - 12 years

☐ > 12 years

13. In which of the following cases, do you prescribe nasal corticosteroids to children with PCD at you centre?

☐ Only during exacerbations

☐ Only during routine care

☐ During both: routine care and exacerbations

*The following question: 14, will appear only if the answers in question 13 are: "Only during routine care", "During both routine care and exacerbations")*

14. Please describe in detail the three most important factors (e.g. specific symptoms or signs seen during examination) that make you prescribe nasal corticosteroids in children with PCD at your centre, in **routine care**.

*(Free text answer)*

*The following question: 15, will appear only if the answers in question:13 are: "Only during routine care", "During both routine care and exacerbations")*

15. During routine care what is the usual therapeutic scheme of nasal corticosteroids you prescribe to children with PCD at your centre?

*(Free text answer– Please include details such as the daily dose, frequency, duration of treatment scheme and any other relevant instructions)*

16. Are there any factors that make you shorten the usual duration or daily dose of nasal corticosteroids treatment to children with PCD at your centre?

*(Free text answer - Please describe any clinical or patient-related factors that influence your decision to shorten the usual treatment duration/ daily dose.)*

17. Are there any factors that make you increase the usual duration or daily dose of nasal corticosteroids treatment to children with PCD at your centre?

*(Free text answer - Please describe any clinical or patient-related factors that influence your decision to increase usual treatment duration/ daily dose.)*

*The following question: 18, will appear only if the answers in question:13 are: “Only during exacerbations”, “During both routine care and exacerbations”)*

18. During exacerbations, what is the usual therapeutic scheme of nasal corticosteroids you prescribe to children with PCD at your centre?

*(Free text answer – Please include details such as the daily dose, frequency, duration of treatment scheme and any other relevant instructions)*

19. Do you have any additional comments or thoughts on the prescription of nasal corticosteroids to children with PCD?

*(Free text answer– Please include any thoughts on clinical experience, challenges, preferences, or uncertainties related to their use.)*

### Nasal rinsing prescription in adults

20. How often do you prescribe nasal saline irrigation to adults with PCD at your centre?

☐ Rarely (in less than 10% of patients)

☐ In some cases (in about 10-40% of patients)

☐ In about half of the cases (in about 50% of patients)

☐ In the majority of cases (in about 60-90% of patients)

☐ Nearly in all cases (in more than 90% of patients)

21. In which of the following cases, do you prescribe nasal saline irrigation to adults with PCD at your centre?

☐ Only during exacerbations

☐ Only during routine care

☐ During both: routine care and exacerbations

*(The following question:22, will appear only if the answer in question:21 is: “During both routine care and exacerbations” or “Online during routine care”)*

22. During **routine care**, what percentage of saline solution do you usually prescribe for nasal saline irrigation to adults with PCD at your centre? (Select **all** that apply.)

☐ Hypotonic saline solution (less than 0.9 %)

☐ Isotonic saline solution (0.9 %)

☐ Hypertonic saline solution (2-5 %, usually 3.5%)

☐ Highly hypertonic saline solution (>6%)

*The following question: 23, will appear only if the answers in question:21 are: “Only during exacerbations”, “During both routine care and exacerbations”)*

23. During **exacerbations**, what percentage of saline solution do you usually prescribe for nasal saline irrigation to adults with PCD at your centre? (Select **all** that apply.)

☐ Hypotonic saline solution (less than 0.9 %)

☐ Isotonic saline solution (0.9 %)

☐ Hypertonic saline solution (2-5 %, usually 3.5%)

☐ Highly hypertonic saline solution (>6%)

*The following question: 24, will appear only if the answers in any of the questions:22, 23 are: “*Hypertonic saline solution*”, “*Highly hypertonic saline solution*”)*

24. Please describe in detail the **main reason(s)** to prescribe hypertonic saline solution instead of isotonic saline solution to adults with PCD at your centre.

*(Free text answer)*

25. What is the usual combination of volume and pressure of saline do you usually prescribe for nasal irrigation to adults with PCD at your centre?

☐ Low-volume and low-pressure devices (nasal drops and sprays)

☐ Low-volume and high-pressure devices (pressurized spray and syringes)

☐ High-volume and low-pressure devices (pots and nebulizers)

☐ High-volume and high-pressure devices (squeeze bottles, bulb syringes, and powered irrigation devices)

☐ I do not usually specify the volume and pressure

26. What factors do you consider for prescribing nasal saline irrigation to adults with PCD at your centre?

*(Free text answer – Please describe any clinical or patient-related factors that make you prescribe nasal saline irrigation treatment.)*

27. Do you have any additional comments or thoughts on the prescription of nasal saline irrigation to adults with PCD?

*(Free text answer – Please include any thoughts on clinical experience, challenges, preferences, or uncertainties related to their use.)*

### Nasal rinsing prescription in children

28. How often do you prescribe nasal saline irrigation to children with PCD at your centre ?

☐ Rarely (in less than 10% of patients)

☐ In some cases (in about 10-40% of patients)

☐ In about half of the cases (in about 50% of patients)

☐ In the majority of cases (in about 60-90% of patients)

☐ Nearly in all cases (in more than 90% of patients)

29. From what age do you prescribe nasal saline irrigation to children with PCD at your centre?

☐ < 3 years

☐ 3 – 5 years

☐ 6 - 12 years

☐ > 12 years

30. In which of the following cases, do you prescribe nasal saline irrigation to children with PCD at you centre?

☐ Only during  exacerbations

☐ Only during routine care

☐ During both routine care and exacerbations

*(The following question: 31, will appear only if the answer in question:30 is: "During both routine care and exacerbations" or "Online during routine care")*

31. During **routine care** what percentage of saline solution do you usually prescribe for nasal saline irrigation to children with PCD at your centre? (Select **all** that apply.)

☐ Hypotonic saline solution (less than 0.9 %)

☐ Isotonic saline solution (0.9 %)

☐ Hypertonic saline solution (2-5 %, usually 3.5%)

☐ Highly hypertonic saline solution (>6%)

*The following question: 32, will appear only if the answers in question: 30 are: "Only during exacerbations", "During both routine care and exacerbations")*

32. During **exacerbations** what percentage of saline solution do you usually prescribe for nasal saline irrigation to children with PCD at your centre? (select **all** that apply)

☐ Hypotonic saline solution (less than 0.9 %)

☐ Isotonic saline solution (0.9 %)

☐ Hypertonic saline solution (2-5 %, usually 3.5%)

☐ Highly hypertonic saline solution (>6%)

*The following question: 33, will appear only if the answers in any of the questions:31, 32 are: "*Hypertonic saline solution*", "*Highly hypertonic saline solution*")*

33. Please describe in detail the **main reason(s)** to prescribe hypertonic saline solution instead of isotonic saline solution to children with PCD at your centre.

*(Free text answer)*

34. What is the usual combination of volume and pressure of saline do you usually prescribe for nasal irrigation to children ≥ 6 years old with PCD at your centre?

☐ Low-volume and low-pressure devices (nasal drops and sprays)

☐ Low-volume and high-pressure devices (pressurized spray and syringes)

☐ High-volume and low-pressure devices (pots and nebulizers)

☐ High-volume and high-pressure devices (squeeze bottles, bulb syringes, and powered irrigation devices)

☐ I do not usually specify the volume and pressure

35. What **factors** do you consider for prescribing nasal saline irrigation to children with PCD at your centre?

*(Free text answer – Please describe any clinical or patient-related factors that make you prescribe nasal saline irrigation treatment.)*

36. Do you have any **additional comments or thoughts** on the prescription of nasal saline irrigation to children with PCD?

*(Free text answer– Please include any thoughts on clinical experience, challenges, preferences, or uncertainties related to their use.)*

### Prescription of other nasal/otologic medications and non-surgical interventions in adults

37. Do you routinely prescribe any of the following nasal treatments to adults with PCD at your centre ? (Select **all** that apply.)

☐ Nasal decongestants (e.g., xylometazoline, oxymetazoline)

☐ Nasal antihistamine sprays

☐ None of the above

38. Do you routinely prescribe any of the following otologic treatments **to adults** with PCD at your centre? (Select **all** that apply)

☐ Ototopical ear drops

☐ Ototopical corticosteroids drops

☐ Use of auto-inflation device to help with eustachian tube dysfunction (e.g. Otovent)

☐ None of the above

### Prescription of other nasal/otologic medications and non-surgical interventions in children

39. Do you routinely prescribe any of the following nasal treatments to children with PCD at your centre? (Select **all** that apply.)

☐ Nasal decongestants (e.g., xylometazoline, oxymetazoline)

☐ Nasal antihistamine sprays

☐ None of the above

40. Do you routinely prescribe any of the following otologic treatments **to children** with PCD at your centre? (Select **all** that apply.)

☐ Ototopical ear drops

☐ Ototopical corticosteroids drops

☐ Use of auto-inflation device to help with eustachian tube dysfunction (e.g. Otovent)

☐ None of the above

### Eradication treatment of upper airways in adults

*Eradication treatment of upper airways is the complete elimination of the presence and growth of microorganisms (such as bacteria or fungi) on mucosal surface of upper airways.*

41. If colonization of the upper airway is detected in adults with PCD at your centre, do you initiate an eradication protocol?

*Colonization of the upper airway is the presence and growth of microorganisms (such as bacteria or fungi) on mucosal surface of upper airway without necessarily causing symptoms or active infection.*

☐ Yes, in all cases

☐ Yes, in some cases

☐ No, we do not treat colonization of upper airway system unless there is an indication to treat parallel lower airway colonisation

*(The following questions: will appear only if the answer in question:41 is: "Yes, in all cases" or "Yes, in some cases")*

42. Which are the **three most important factors** that make you apply eradication treatment in adults with PCD at your centre?

*(Free text answer - Please describe any clinical or patient-related factors that make you suggest an eradication treatment.)*

43. Do you have **any additional comments or thoughts on** the suggestion of eradication treatment in adults with PCD?

*(Free text answer – Please include any thoughts on clinical experience, challenges, preferences, or uncertainties related to their use.)*

### Eradication treatment of upper airways in children

*Eradication treatment of upper airways is the complete elimination of the presence and growth of microorganisms (such as bacteria or fungi) on mucosal surface of upper airways.*

44. If colonization of the upper airway is detected in children with PCD at your centre, do you initiate an eradication protocol?

*Colonization of the upper airway is the presence and growth of microorganisms (such as bacteria or fungi) on mucosal surface of upper airway without necessarily causing symptoms or active infection.*

☐ Yes, in all cases

☐ Yes, in some cases

☐ No, we do not treat colonization of upper airway system unless there is an indication to treat parallel lower airway colonisation.

*(The following questions: will appear only if the answer in question:44 is: "Yes, in all cases" or "Yes, in some cases")*

45. Which are the **three most important factors** that make you apply eradication treatment in children with PCD at your centre?

*(Free text answer - Please describe any clinical or patient-related factors that make you suggest an eradication treatment. )*

46. Do you have any additional comments or thoughts on the suggestion of eradication treatment in children with PCD?

*(Free text answer – Please include any thoughts on clinical experience, challenges, preferences, or uncertainties related to their use.)*

### Surgical practices

### Functional Endoscopic Sinus Surgery (FESS)

47. How often do you recommend Functional Endoscopic Sinus Surgery (FESS) to adults with PCD at your centre?

☐ Rarely (in less than 10% of patients)

☐ In some cases (in about 10-40% of patients)

☐ In about half of the cases (in about 50% of patients)

☐ In the majority of cases (in about 60-90% of patients)

☐ Nearly in all cases (in more than 90% of patients)

48. What are the **main factors** to recommend Functional Endoscopic Sinus Surgery (FESS) to adults with PCD at your centre?

*(Free text answer -Please describe any clinical or patient-related factors that make you suggest an eradication treatment.)*

49. How often do you recommend Functional Endoscopic Sinus Surgery (FESS) **to children** with PCD at your centre?

☐ Rarely (in less than 10% of patients)

☐ In some cases (in about 10-40% of patients)

☐ In about half of the cases (in about 50% of patients)

☐ In the majority of cases (in about 60-90% of patients)

☐ Nearly in all cases (in more than 90% of patients)

50. What are the main factors to recommend Functional Endoscopic Sinus Surgery (FESS) to children with PCD at your centre?

*(Free text answer - Please describe any clinical or patient-related factors that make you suggest an eradication treatment. )*

### Polypectomy

51. How often do you recommend polypectomy to adults with PCD at your centre?

☐ Rarely (in less than 10% of patients)

☐ In some cases (in about 10-40% of patients)

☐ In about half of the cases (in about 50% of patients)

☐ In the majority of cases (in about 60-90% of patients)

☐ Nearly in all cases (in more than 90% of patients)

52. What are the main factors to recommend polypectomy to adults with PCD at your centre?

*(Free text answer - Please describe any clinical or patient-related factors that make you suggest an eradication treatment.)*

53. How often do you recommend polypectomy to children with PCD at your centre?

☐ Rarely (in less than 10% of patients)

☐ In some cases (in about 10-40% of patients)

☐ In about half of the cases (in about 50% of patients)

☐ In the majority of cases (in about 60-90% of patients)

☐ Nearly in all cases (in more than 90% of patients)

54. What are the main factors to recommend polypectomy to children with PCD at your centre?

*(Free text answer - Please describe any clinical or patient-related factors that make you suggest an eradication treatment.)*

### Myringotomy with ventilation tube insertion

55. How often do you recommend myringotomy with ventilation tube insertion to adults with PCD at your centre?

☐ Rarely (in less than 10% of patients)

☐ In some cases (in about 10-40% of patients)

☐ In about half of the cases (in about 50% of patients)

☐ In the majority of cases (in about 60-90% of patients)

☐ Nearly in all cases (in more than 90% of patients)

56. What are the main factors to recommend myringotomy with ventilation tube insertion to adults with PCD at your centre?

*(Free text answer -Please describe any clinical or patient-related factors that make you suggest an eradication treatment. )*

57. How often do you recommend myringotomy with ventilation tube insertion to children with PCD at your centre?

☐ Rarely (in less than 10% of patients)

☐ In some cases (in about 10-40% of patients)

☐ In about half of the cases (in about 50% of patients)

☐ In the majority of cases (in about 60-90% of patients)

☐ Nearly in all cases (in more than 90% of patients)

58. What are the main factors to recommend myringotomy with ventilation tube insertion to **children** with PCD at your centre?

*(Free text answer - Please describe any clinical or patient-related factors that make you suggest an eradication treatment. )*

59. Which **other surgical interventions** related to the upper airways do you usually recommend **to adults** with PCD at your centre?

*(Free text answer)*

60. Which **other surgical interventions** related to the upper airways do you usually recommend **to children** with PCD at your centre?

*(Free text answer)*

61. Do you have any additional comments or thoughts on surgical practices of upper airway disease in **patients** with PCD?

(*Free text answer – Please include any thoughts on clinical experience, challenges, preferences, or uncertainties related to their performance.)*

Thank you for answering this survey! Your answers will provide valuable insights into the management of upper airway disease in patients with PCD.

### Supplementary Results

**Upper airway disease in primary ciliary dyskinesia: Clinical management and factors influencing decision-making, a multicentre analysis, Vasiliki Gkatzou, Alexis Campos, Nena Karavasiloglou, …, on behalf of the EPIC-PCD team, Myrofora Goutaki**

### Supplementary results outline

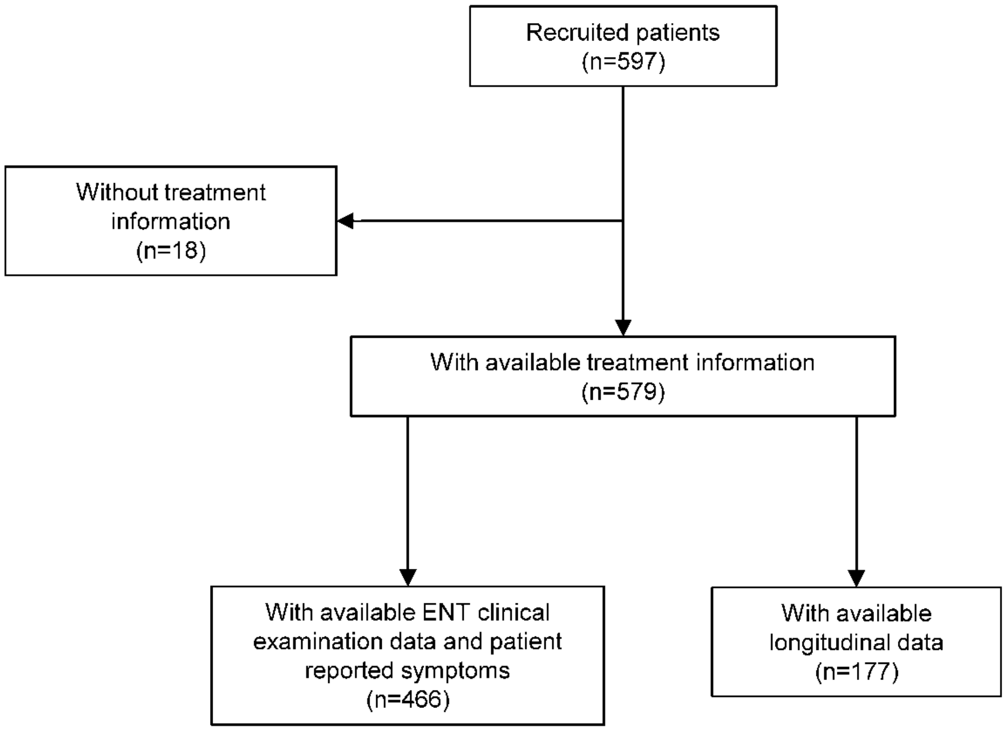

**Figure S1.** Flowchart of the EPIC-PCD study population

*EPIC-PCD: Ear-nose throat Prospective International Cohort of patients with Primary Ciliary Dyskinesia*

**Table S1. Sinonasal examination results of EPIC-PCD participants, overall (n=466) and by age group**

| | Total | Age 0-6 y | Age 7-12 y | Age 13-17 y | Age 18-30 y | Age 31-50 y | Age >51 y |
|---|---|---|---|---|---|---|---|
| **Participants** | 466 (100) | 61 (100) | 113 (100) | 109 (100) | 93 (100) | 58 (100) | 32 (100) |
| **Blocked nose** | 182 (39) | 16 (26) | 38 (34) | 41 (38) | 41 (44) | 32 (55) | 14 (44) |
| Not recorded | 5 (1) | 3 (5) | 1 (1) | 1 (1) | 0 (0) | 0 (0) | 0 (0) |
| **Nasal discharge** | 354 (76) | 40 (66) | 83 (74) | 81 (74) | 78 (84) | 46 (79) | 26 (81) |
| Type | | | | | | | |
| Serous | 127 (27) | 20 (33) | 34 (30) | 26 (24) | 23 (25) | 16 (28) | 8 (25) |
| Sero-mucous | 152 (33) | 14 (23) | 37 (33) | 33 (30) | 38 (41) | 20 (35) | 10 (31) |
| Muco-purulent | 69 (15) | 5 (8) | 12 (11) | 19 (17) | 17 (18) | 9 (16) | 7 (22) |
| Mixed with blood | 3 (1) | 1 (2) | 0 (0) | 1 (1) | 0 (0) | 0 (0) | 1 (3) |
| Not recorded | 3 (1) | 0 (0) | 0 (0) | 2 (2) | 0 (0) | 1 (2) | 0 (0) |
| **Abnormal nasal mucosa** | 207 (44) | 17 (28) | 46 (41) | 51 (47) | 42 (45) | 30 (52) | 21 (66) |
| Not recorded | 19 (4) | 8 (13) | 6 (5) | 3 (3) | 2 (2) | 0 (0) | 0 (0) |
| **Nasal polyps** | 59 (13) | 2 (3) | 5 (4) | 17 (16) | 15 (16) | 15 (26) | 5 (16) |
| Not recorded | 50 (11) | 22 (36) | 18 (16) | 5 (5) | 3 (3) | 1 (2) | 1 (3) |
| **Nasal turbinates** | | | | | | | |
| Normal | 276 (59) | 40 (66) | 64 (57) | 61 (56) | 64 (69) | 28 (48) | 19 (60) |
| Hypertrophy | 158 (34) | 12 (20) | 42 (37) | 45 (41) | 27 (29) | 24 (41) | 8 (25) |
| Atrophy | 5 (1) | 0 (0) | 0 (0) | 0 (0) | 0 (0) | 4 (7) | 1 (3) |
| Not recorded | 27 (6) | 9 (15) | 7 (6) | 3 (3) | 2 (2) | 2 (3) | 4 (13) |
| **Deviated septum** | 131 (28) | 4 (7) | 24 (21) | 34 (31) | 37 (40) | 19 (33) | 13 (41) |
| Not recorded | 33 (7) | 13 (21) | 10 (9) | 4 (4) | 5 (5) | 1 (2) | 0 (0) |
| **Facial pain/ sensitivity** | 53 (11) | 0 (0) | 5 (4) | `12 (11) | 11 (12) | 13 (22) | 12 (38) |
| Not recorded | 27 (6) | 13 (21) | 3 (3) | 5 (5) | 4 (4) | 2 (3) | 0 (0) |

*EPIC-PCD: Ear-nose throat Prospective International Cohort of patients with Primary Ciliary Dyskinesia; PCD: Primary Ciliary Dyskinesia; y: years.*
*Data is presented as counts (percentages).*

### Table S2. Otologic examination results of EPIC-PCD participants, overall (n=466) and by age group

| | Total | Age 0-6 y | Age 7-12 y | Age 13-17 y | Age 18-30 y | Age 31-50 y | Age >51 y |
|---|---|---|---|---|---|---|---|
| **Participants** | 466 (100) | 61 (100) | 113 (100) | 109 (100) | 93 (100) | 58 (100) | 32 (100) |
| **Ear discharge** | 40 (9) | 2 (3) | 13 (12) | 16 (15) | 4 (4) | 2 (3) | 3 (9) |
| Not recorded | 11 (2) | 5 (8) | 3 (3) | 0 (0) | 1 (1) | 2 (3) | 0 (0) |
| **Tympanic perforation** | 35 (8) | 0 (0) | 7 (6) | 16 (15) | 8 (9) | 3 (5) | 1 (3) |
| Not recorded | 16 (3) | 7 (12) | 5 (4) | 2 (2) | 1 (1) | 1 (2) | 0 (0) |
| **Retracted tympanic membrane** | 64 (14) | 2 (3) | 10 (9) | 17 (16) | 16 (17) | 9 (16) | 10 (31) |
| Not recorded | 25 (5) | 9 (15) | 5 (4) | 6 (6) | 3 (3) | 2 (3) | 0 (0) |
| **Acute otitis media** | 4 (1) | 0 (0) | 3 (3) | 0 (0) | 1 (1) | 0 (0) | 0 (0) |
| Not recorded | 20 (4) | 7 (12) | 5 (4) | 5 (5) | 1 (1) | 2 (3) | 0 (0) |
| **Otitis media with effusion** | 154 (33) | 27 (44) | 46 (41) | 41 (38) | 20 (22) | 8 (14) | 12 (38) |
| Not recorded | 23 (5) | 8 (13) | 7 (6) | 3 (3) | 3 (3) | 2 (3) | 0 (0) |
| **Tympanic sclerosis** | 96 (21) | 1 (2) | 12 (11) | 19 (17) | 28 (30) | 20 (34) | 16 (50) |
| Not recorded | 38 (8) | 12 (20) | 11 (10) | 9 (8) | 3 (3) | 3 (5) | 0 (0) |
| **Tympanometry performed** | 274 (59) | 24 (39) | 62 (55) | 65 (60) | 59 (63) | 38 (66) | 26 (81) |
| **Tympanogram ¶** | | | | | | | |
| Type A \| Type A | 66 (14) | 2 (3) | 14 (12) | 17 (16) | 24 (26) | 7 (12) | 2 (6) |
| Type AD \| Type A | 2 (0) | 1 (2) | 0 (0) | 0 (0) | 0 (0) | 1 (2) | 0 (0) |
| Type AS \| Type A | 6 (1) | 0 (0) | 0 (0) | 3 (3) | 3 (3) | 0 (0) | 0 (0) |
| Type AD \| Type AD | 1 (0) | 0 (0) | 0 (0) | 1 (1) | 0 (0) | 0 (0) | 0 (0) |
| Type AS \| Type AS | 21 (5) | 0 (0) | 4 () | 6 (6) | 3 (3) | 6 (10) | 2 (6) |
| Type AS \| Type AD | 1 (0) | 0 (0) | 0 (0) | 0 (0) | 1 (1) | 0 (0) | 0 (0) |
| Type B \| Type A | 9 (2) | 2 (3) | 3 (3) | 3 (3) | 1 (1) | 0 (0) | 0 (0) |
| Type B \| Type AS | 6 (1) | 1 (2) | 2 (2) | 0 (0) | 2 (2) | 0 (0) | 1 (3) |
| Type B \| Type B | 117 (25) | 11 (18) | 32 (28) | 27 (25) | 15 (16) | 15 (26) | 17 (53) |
| Type C \| Type A | 7 (2) | 0 (0) | 1 (1) | 3 (3) | 2 (2) | 1 (2) | 0 (0) |
| Type C \| Type B | 11 (2) | 1 (2) | 3 (3) | 2 (2) | 2 (2) | 1 (2) | 2 (6) |
| Type C \| Type AD | 1 (0) | 0 (0) | 0 (0) | 0 (0) | 0 (0) | 1 (2) | 0 (0) |
| Type C \| Type AS | 2 (0) | 0 (0) | 0 (0) | 0 (0) | 0 (0) | 1 (2) | 1 (3) |
| Type C \| Type C | 15 (3) | 5 (8) | 2 (2) | 2 (2) | 4 (4) | 2 (3) | 0 (0) |
| Not recorded | 9 (2) | 1 (2) | 1 (1) | 1 (1) | 2 (2) | 3 (5) | 1 (3) |
| **Grommets** | 47 (10) | 4 (7) | 11 (10) | 15 (14) | 9 (10) | 6 (10) | 2 (6) |
| Not recorded | 39 (8) | 11 (18) | 10 (9) | 9 (8) | 1 (1) | 2 (3) | 6 (19) |

| | | | | | | | |
|---|---|---|---|---|---|---|---|
| **Audiometry performed** | 317 (68) | 25 (41) | 73 (65) | 69 (63) | 69 (74) | 52 (90) | 29 (91) |
| Pure tone | 231 (50) | 14 (23) | 54 (48) | 57 (52) | 51 (55) | 33 (57) | 22 (69) |
| Vocal | 7 (2) | 2 (3) | 3 (3) | 1 (1) | 1 (1) | 0 (0) | 0 (0) |
| Bone conduction | 7 (2) | 0 (0) | 2 (2) | 1 (1) | 3 (3) | 1 (2) | 0 (0) |
| Other | 50 (10) | 8 (13) | 7 (6) | 5 (5) | 10 (11) | 16 (28) | 4 (13) |
| Not recorded | 22 (4) | 1 (2) | 7 (6) | 5 (5) | 4 (4) | 2 (3) | 3 (9) |
| **Audiometry results #** | | | | | | | |
| Normal\| Normal | 173 (37) | 10 (16) | 46 (41) | 40 (37) | 47 (51) | 27 (47) | 3 (9) |
| Normal \| Mild | 32 (7) | 1 (2) | 8 (7) | 12 (11) | 6 (6) | 4 (7) | 1 (3) |
| Normal \| Moderate | 1 (0) | 0 (0) | 0 (0) | 0 (0) | 1 (1) | 0 (0) | 0 (0) |
| Mild\| Mild | 70 (15) | 11 (18) | 13 (12) | 10 (9) | 11 (12) | 15 (26) | 10 (31) |
| Mild \| Moderate | 16 (3) | 0 (0) | 3 (3) | 3 (3) | 3 (3) | 5 (9) | 2 (6) |
| Mild \| Severe | 2 (0) | 0 (0) | 0 (0) | 1 (1) | 0 (0) | 0 (0) | 1 (3) |
| Moderate\| Moderate | 12 (3) | 2 (3) | 1 (1) | 0 (0) | 0 (0) | 0 (0) | 9 (28) |
| Moderate \| Severe | 3 (1) | 0 (0) | 0 (0) | 1 (1) | 0 (0) | 1 (2) | 1 (3) |
| Moderate\| Profound | 1 (0) | 0 (0) | 0 (0) | 0 (0) | 1 (1) | 0 (0) | 0 (0) |
| Severe\| Severe | 3 (1) | 0 (0) | 0 (0) | 1 (1) | 0 (0) | 0 (0) | 2 (6) |
| Profound\| Profound | 1 (0) | 0 (0) | 1 (1) | 0 (0) | 0 (0) | 0 (0) | 0 (0) |
| Not recorded | 3 (1) | 1 (2) | 1 (1) | 1 (1) | 0 (0) | 0 (0) | 0 (0) |
| **Hearing aids** | 42 (9) | 5 (8) | 13 (12) | 10 (9) | 1 (1) | 6 (10) | 7 (22) |

*EPIC-PCD: Ear-nose throat Prospective International Cohort of patients with Primary Ciliary Dyskinesia; PCD: Primary Ciliary Dyskinesia; y: years.*
*¶Tympanogram types (A, AD, AS, C, and C) follow the Jerger classification; #Hearing loss classification based on the World Health Organization (WHO) grading system: normal hearing, 0–25 dB; mild hearing loss, 26–40 dB; moderate hearing loss, 41–60 dB; severe hearing loss, 61–80 dB; profound hearing loss, >80 dB.*
*Data is presented as counts (percentages).*

**Table S3. Upper respiratory symptoms of past 3 months reported by EPIC-PCD participants, overall and by age group (n=466)**

| | Total | Age 0-6 y | Age 7-12 y | Age 13-17 y | Age 18-30 y | Age 31-50 y | Age >51 y |
|---|---|---|---|---|---|---|---|
| **Participants** | 466 (100) | 61 (100) | 113 (100) | 109 (100) | 93 (100) | 58 (100) | 32 (100) |
| **Nasal symptoms** | | | | | | | |
| Frequent | 242 (52) | 33 (54) | 55 (49) | 44 (40) | 51 (55) | 35 (60.5) | 24 (75) |
| Infrequent | 216 (46) | 26 (43) | 58 (51) | 64 (59) | 39 (42) | 21 (36) | 8 (25) |
| Not reported | 8 (2) | 2 (3) | 0 (0) | 1 (1) | 3 (3) | 2 (3.5) | 0 (0) |
| **Type of nasal symptoms*** | | | | | | | |
| Blocked nose | 283 (61) | 22 (36) | 69 (61) | 72 (66) | 56 (60) | 42 (72) | 22 (69) |
| Rhinorrhoea | 297 (64) | 49 (80) | 67 (59) | 60 (55) | 60 (65) | 38 (66) | 23 (72) |
| Sneezing | 91 (20) | 8 (13) | 18 (16) | 17 (16) | 22 (24) | 14 (24) | 12 (38) |
| Anosmia/Hyposmia | 49 (11) | 0 (0) | 2 (2) | 9 (8) | 8 (9) | 15 (26) | 15 (47) |
| **Headache** | | | | | | | |
| Frequent | 55 (12) | 1 (2) | 9 (8) | 13 (12) | 11 (12) | 11 (19) | 10 (31) |
| Infrequent | 374 (80) | 33 (54) | 103 (91) | 93 (85) | 79 (85) | 45 (78) | 21 (66) |
| Not reported | 37 (8) | 27 (44) | 1 (1) | 3 (3) | 3 (3) | 2 (3) | 1 (3) |
| **Ear pain** | | | | | | | |
| Frequent | 59 (13) | 2 (3) | 9 (8) | 13 (12) | 13 (14) | 12 (21) | 10 (31) |
| Infrequent | 376 (81) | 40 (66) | 101 (89) | 90 (83) | 78 (84) | 46 (79) | 21 (66) |
| Not reported | 31 (7) | 19 (31) | 3 (3) | 6 (6) | 2 (2) | 0 (0) | 1 (3) |
| **Ear discharge** | | | | | | | |
| Frequent | 24 (5) | 0 (0) | 4 (3.5) | 7 (6) | 5 (5) | 5 (9) | 3 (9) |
| Infrequent | 428 (92) | 60 (98) | 108 (95.5) | 99 (91) | 86 (93) | 50 (86) | 25 (78) |
| Not reported | 14 (3) | 1 (2) | 1 (1) | 3 (3) | 2 (2) | 3 (5) | 4 (13) |
| **Hearing problems** | | | | | | | |
| Frequent | 94 (20) | 9 (15) | 17 (15) | 14 (13) | 15 (16) | 19 (33) | 20 (63) |
| Infrequent | 339 (73) | 40 (65.5) | 89 (79) | 88 (81) | 75 (81) | 37 (64) | 10 (31) |
| Not reported | 33 (7) | 12 (19.5) | 7 (6) | 7 (6) | 3 (3) | 2 (3) | 2 (6) |
| **Snoring** | | | | | | | |
| Frequent | 49 (10.5) | 6 (10) | 10 (9) | 7 (6) | 8 (9) | 11 (19) | 7 (22) |
| Infrequent | 354 (76) | 54 (88.5) | 90 (79.5) | 89 (82) | 66 (71) | 38 (65.5) | 17 (53) |
| Not reported | 63 (13.5) | 1 (1.5) | 13 (11.5) | 13 (12) | 19 (20) | 9 (15.5) | 8 (25) |

*EPIC-PCD: Ear-nose throat Prospective International Cohort of patients with Primary Ciliary Dyskinesia; PCD: Primary Ciliary Dyskinesia*

*Data is presented as counts (percentages) for EPIC-PCD participants with available information on upper airway disease management practices, clinical examination signs and patient reported data, recorded on the same date or within 14 days. Symptoms were reported using a five-point frequency scale (never/rarely/sometimes/often/daily). These were recoded into two categories: frequent (often, daily) and infrequent (never, rarely, sometimes). *Categories are not mutually exclusive.*

### ENT-specialists' survey results

One ear-nose throat (ENT) specialist from each EPIC-PCD study participating centre completed the survey, in total 14 specialists, including 3 treating only children and 11 treating both children and adults. The 15th participating study centre joined the study after circulation of this survey thus no specialist participated. Specialists managing all age groups completed the questionnaire separately for paediatric and adult care. Nasal rinsing was widely used across centres. Most specialists reported prescribing nasal rinsing routinely in ≥60% of patients (children: 10/14, 71%; adults: 8/11, 73%). Only a minority reported rare use (<10% of patients) (children: 1/14, 7%; adults: 0/11). Prescription patterns were similar between age groups, with most specialists prescribing nasal rinsing both during routine care and exacerbations (children: 10/14, 71%; adults: 8/11, 73%). In contrast, nasal corticosteroid use was more heterogeneous. In children, 6 specialists (43%) reported rare use (<10% of patients), whereas only 1 (7%) prescribed them in ≥60% of patients. In adults, routine use (≥60% of patients) was more frequent (3/11, 27%), although two ENT specialists (18%) reported rare use. Nasal corticosteroids were most commonly prescribed during exacerbations (children: 7/14, 50%; adults: 5/11, 46%) or both during routine care and exacerbations (children: 6/14, 43%; adults: 6/11, 55%). Exclusive prescription of corticosteroids during routine care use was uncommon (children: 1/14, 7%; adults: 0/11).

The use of surgical practices varied, depending on the type of surgery. Most specialists reported performing polypectomy in <10% of patients (children: 14/14, 100%; adults: 9/11, 82%). Greater variability was observed for Functional Endoscopic Sinus Surgery (FESS). In children, 8 specialists (57%) reported performing FESS in <10% of patients, whereas 5 (36%) reported use in 10–40% of patients. In adults, frequency of use was higher, with 5 specialists 11 (45%) reporting rare use (<10%) and 5 (45%) reporting use in 10–50% of patients. Similarly, grommet insertion showed heterogeneity across centres, with use ranging from rare to more frequent categories in both age groups. Among ENT specialists treating children, 8 (57%) reported using grommets occasionally (in 10–50% of patients), 5 (36%) rarely (in <10% of patients), and only one (7%) routinely (in ≥60% of patients). Among those treating adults, the majority reported using grommets rarely (8/11, 67%), with 3 (25%) reporting occasional use and one (8%) routine use.

For medical treatments, decision-making differed between nasal corticosteroids and nasal rinsing. ENT specialists prescribed nasal corticosteroids mainly based on symptom burden and structural findings. Nasal obstruction, nasal discharge, and nasal polyps were the key drivers for nasal corticosteroids prescription in both children and adults. In contrast, specialists viewed nasal rinsing as a fundamental treatment for upper airway disease. In children, the presence of secretions was an important driver to prescribe nasal rinsing. In both children and adults, practical factors, namely ability to use the device, patient training, and reimbursement issues, strongly influenced the prescription of nasal rinsing. For surgical treatments, FESS decisions were mainly driven by structural severity, especially extensive nasal polyposis. Specialists also considered severe clinical phenotype, facial pain or headaches, and failure of medical therapy. In children, age influenced decisions, and FESS was generally avoided below 12 years. Polypectomy was primarily driven by structural severity, such as massive or high-grade polyps and total obstruction. Specialists also considered disease severity, suspected neoplasm, exacerbations in adults, patient complaints, and failure of medical therapy. Age was an important driver that influenced decisions, with polypectomy also usually avoided below 12 years. Several specialists stated that isolated polypectomy should be avoided and combined with FESS. For grommets insertion, the main drivers were structural ear disease and significant hearing loss. Specialists considered severe chronic otitis, persistent effusion, and hearing loss greater than 40 db. In children, developmental factors such as speech delay and learning problems played an additional role. Failure of medical therapy and inability to tolerate hearing aids also influenced decisions. Some specialists considered overall disease context, including lower airway disease status (Table 4).

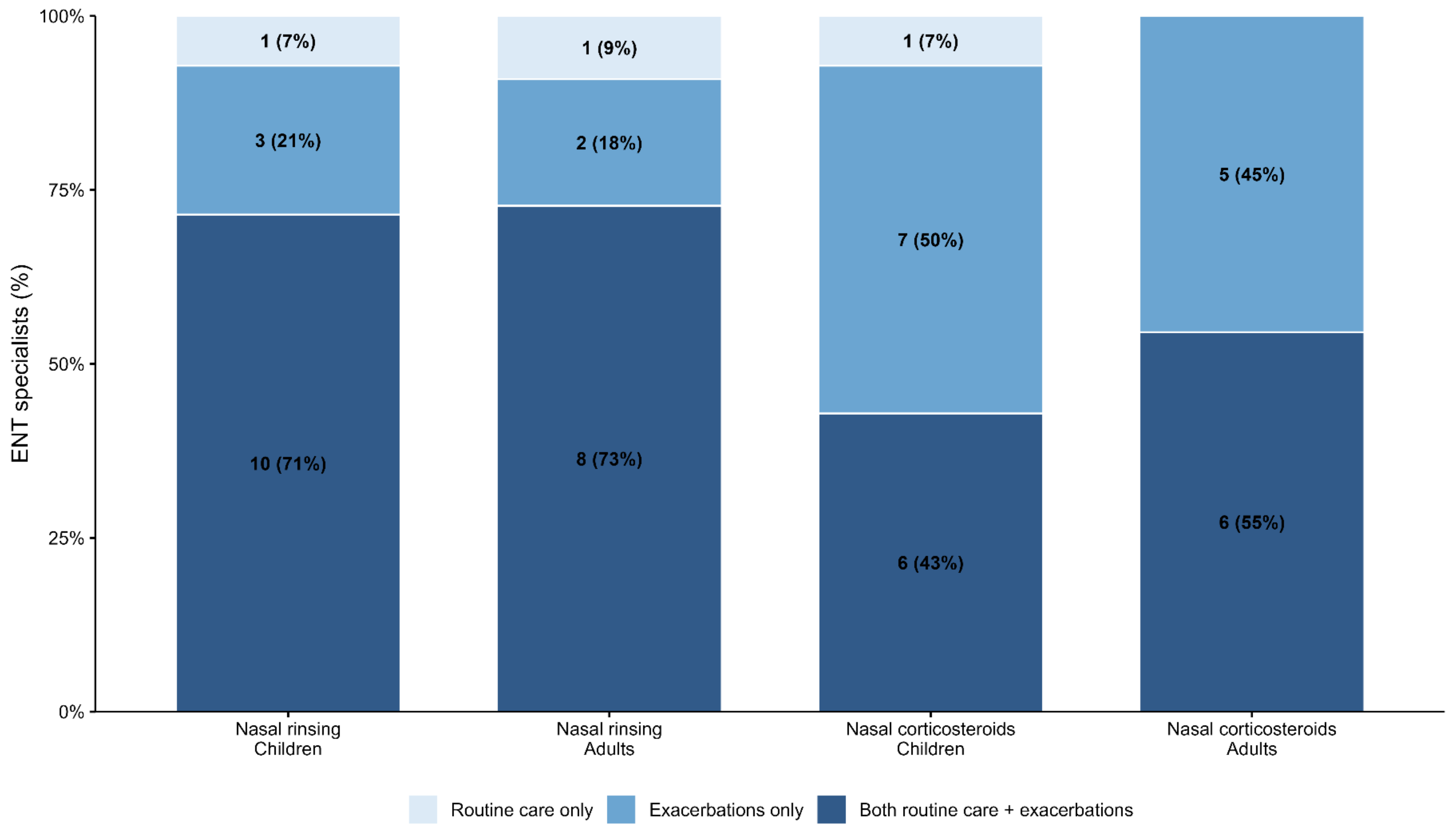

**Figure S2.** Frequency of recommendation of upper airway disease management practices – Results from the ENT-specialists survey

*ENT: Ear-Nose-Throat. Bars represent the proportion of ENT specialists prescribing each management practice during routine care only, during exacerbations only, or during both routine care and exacerbations. Percentages are calculated within each age group, as 3 specialists were treating only children and 11 were treating both children and adults (Children: n = 14; Adults: n = 11). ENT specialists treating both children and adults completed the survey separately for paediatric and adult care. Values inside bars represent the count of respondents and the corresponding percentage.*
